# A Longitudinal and Reproducible Anti-coactivation Pattern Between the Cerebellum and the Ventral Tegmental Area Relates to Apathy in Schizophrenia

**DOI:** 10.1101/2024.07.11.24310281

**Authors:** Farnaz Delavari, Jade Awada, Thomas A. W. Bolton, Fares Alouf, Fabien Carruzzo, Noemie Kuenzi, Mariia Kaliuzhna, Tal Geffen, Teresa Katthagen, Florian Schlagenhauf, Dimitri Van De Ville, Stephan Eliez, Stefan Kaiser, Indrit Bègue

## Abstract

**Background:** Negative symptoms of schizophrenia lack effective treatments. Anomalies in the reward system and cerebellum have been linked to negative symptom The cerebellum modulates reward circuitry via the ventral tegmental area (VTA). The “cognitive dysmetria theory” posits that reduced cerebellar inhibition in schizophrenia may underlie striatal hyperdopaminergia. However, cerebellum-VTA connectivity and its impact on negative symptoms in schizophrenia remains unclear.

**Methods:** From 427 individuals screened, 146 participants were recruited: 90 with schizophrenia (SZ) and 56 healthy controls (HC). At 3 months (T2), 65 individuals (36 SZ, 29 HC) completed follow-up. SZ participants were invited for clinical interviews at 9 months (T3; 33 SZ). After quality check, 105 participants were retained at T1, 41 at T2, and 21 at T3. The validation cohort consisted of 53 individuals (28 SZ, 25 HC). The Brief Negative Symptom Scale was used to quantify negative symptoms. Dynamic functional connectivity of the cerebellum and VTA was analyzed using state-of-the-art coactivation patterns analysis.

**Results:** A reproducible cerebellum-VTA anti-coactivation pattern was found across T1 and T2 (r = 0.98) in bilateral paravermal Crus I/II. Lower anti-coactivation emergence at T1 correlated with worse apathy, particularly asociality and avolition. At T2, lower anti-coactivation persistence related to worse apathy, especially anhedonia, and correlated with worse anhedonia at T3. Similarly, reduced anti-coactivation emergence at T2 linked to worse asociality at T3. In the validation cohort, we replicated the anti-coactivation pattern (r = 0.93) and the correlation of its emergence with apathy, in particular, asociality.

**Conclusion:** Reduced cerebellum-VTA anti-coactivation is a reproducible neural marker of apathy in schizophrenia, highlighting its potential as a target for therapeutic intervention.

## INTRODUCTION

Negative symptoms in schizophrenia – a chronic psychiatric condition affecting up to 1% of the population – are marked by features such as apathy (asociality, anhedonia and avolition) or diminished expression (blunted affect, alogia) (1). Negative symptoms represent a key unmet need in psychiatry due to their profound effect on social functioning, their prevalence across various stages of the disorder and antipsychotic medication resistance (1). Therefore, a better understanding of the underlying mechanisms and pathways involved in negative symptoms in schizophrenia is crucial for advancing treatment options.

Recent conceptualizations of negative symptoms have evolved to map these symptoms onto two dimensions: apathy, manifesting as reductions in goal-directed behavior (avolition), pleasure (anhedonia) and social communication (asociality), and diminished expression in the emotional and verbal domains (blunted affect and alogia, respectively) (2). Emerging evidence highlights a link between negative symptoms, in particular apathy, and dysfunctions in the reward system including the Ventral Tegmental Area (VTA; mesolimbic pathway) (3,4). Parallel to these findings, the cerebellum, traditionally recognized for its role in motor coordination, has been increasingly implicated in reward processing (5), social and emotional learning (6), and the pathophysiology of negative symptoms in schizophrenia (4,7). From early neuroimaging to modern day research, cerebellar vermis structural reduction (8) and altered cerebellar-cortical functional connectivity (9–11) have pointed to a key role of the cerebellum in schizophrenia.

A seminal theory on cerebellar involvement in schizophrenia was conceptualized with the Cognitive Dysmetria Theory (12), given links between forebrain hyperdopaminergia and cerebellar vermis malfunction from reductions in Purkinje cell size and density in post-mortem studies (13). This theory postulated that malfunction of the cerebellar cortex would lead to disinhibition of excitatory projections from the deep nuclei to dopaminergic neurons in the midbrain, resulting in a “hyperdopaminergic state” (14). Consistent with this hypothesis, Daskalakis et al. (2005) examined cerebellar brain inhibition in patients with schizophrenia and found an almost 30% reduction compared to healthy controls (15). Cerebellar brain inhibition refers to the inhibitory tone exerted by the cerebellum on motor neural circuits manifested as a reduction in motor evoked potential when the cerebellum is stimulated 5-7 ms prior to the motor cortex (16). These findings raise the possibility that the cerebellum may be implicated in the negative symptoms pathophysiology via the exertion of modulatory role on the reward system (17), analogous to its established role on motor control. This hypothesis is grounded in several mechanistic insights tying the cerebellum to reward circuitry: 1) activity within the cerebellum during reward anticipation has been consistently observed in human neuroimaging (18) and rodent studies (19); 2) direct monosynaptic connections from the deep cerebellar nuclei to the VTA have been documented in both animal (20–22) and human studies (23–25); 3) the cerebellum exerts an inhibitory tone on the circuit of Papez (24) and its stimulation was found to be associated to ‘pleasure’ and stopping of seizure starting from the hippocampus (25). In addition, the cerebellum’s contribution to the negative symptoms pathophysiology, especially through its effects on the reward system, can be understood by considering its intrinsic role in coordinating and timing of neural processes (5). Thus, in sum, the cerebellum’s capacity to integrate inputs relevant to reward processing and modulate reward-related activities through precise timing and coordination, could underlie the mechanisms behind cerebellar contributions to the negative symptom pathophysiology.

A key role for the cerebellum with high therapeutic potential to address negative symptoms is further supported from recent non-invasive cerebellar neuromodulation evidence, convergently indicating improvement in negative symptoms severity (26). Active (but not sham) cerebellar transcranial magnetic stimulation (TMS) increased resting-state functional connectivity between the vermian and pallidal areas (27), suggesting a functional linkage with regions in the reward circuitry. Overall, this evidence suggests that the cerebellum’s connectivity with the reward system, especially its interactions with the VTA, should provide clinically relevant insights. However, the functional connectivity between the cerebellum and the VTA in the context of negative symptoms remains largely unknown and has not been examined in schizophrenia.

In this research, we set out to investigate for the first time the connectivity between the cerebellum and the VTA across three time points [baseline (T1), at 3 months (T2) and 9 months (T3) after baseline] in a well-characterized cohort of patients with schizophrenia. To strengthen the generalizability of our findings, we employed an additional independently acquired validation cohort. We leveraged a well-established dynamic functional connectivity approach, coactivation pattern (CAP) analysis (28,29), to capture transient neural coactivation events between the cerebellum and VTA during resting-state. We focused on the VTA as the seed region and extracted its spatial patterns of coactivation within the cerebellum. We subsequently derived metrics related to these spatial patterns: emergence (*i.e.*, CAP entry frequency, quantifying the number of times a certain pattern of coactivation occurred from VTA below-threshold activity) and persistence (*i.e.*, CAP duration, quantifying the time in seconds a certain pattern of coactivation persisted upon occurring). We hypothesized that the features of the dynamic functional connectivity patterns between the VTA and the cerebellum during resting-state were linked to schizophrenia clinical manifestations and would represent a functional signature of negative symptoms in schizophrenia over time.

## METHODS

### Data Collection

We systematically collected demographic, clinical, cognitive, behavioral, and neuroimaging data in a main cohort and a validation cohort.

For the main cohort, from 427 individuals screened, we recruited 146 individuals (90 patients with schizophrenia spectrum disorder (SZ) and 56 healthy controls (HC) at baseline, 65 individuals (36 SZ and 29 HC) at 3 months after baseline (T2), and 33 SZ at 9 months after baseline (T3) (refer to the screening process in Table S1 in the Supplemental Materials). Participants were administered a series of interviews and neuroimaging protocols at T1 and T2, and only interviews at T3. The cohort was recruited in the context of two studies approved by the Geneva Ethics Committee in Switzerland (CCER. BASEC ID: 2017-01765 and 2020-02169). The validation cohort was acquired in the context of the SyMoNe study (“Neural and behavioral markers for the motivational negative symptoms of schizophrenia – a longitudinal approach”; trial registration: DRKS00011405).

We recruited clinically stable patients with schizophrenia (*i.e.*, no changes in medication and no hospitalizations in the last 3 months). Patients had confirmed schizophrenia diagnosis via the Mini International Neuropsychiatric Interview (30). Given our focus in obtaining mechanistical insights and in accordance with recent recommendations (1), we excluded patients with secondary sources of negative symptoms (*e.g.*, psychosis, extrapyramidal effects, medication, depression, sedation; see exclusion criteria in Supplemental material). Patients were recruited from community outpatient centers in the University Hospital of Geneva, Switzerland for the main cohort and from outpatient clinics at the Department of Psychiatry and Psychotherapy of Charité – Universitätsmedizin Berlin for the validation cohort (see Figure S1 in Supplemental Materials).

### Clinical psychopathological and cognitive assessment

In both cohorts, all individuals (HC and SZ) underwent a detailed psychopathological and cognitive assessment (see Supplemental Materials for comprehensive description of the assessment). To assess the negative symptoms in patients with schizophrenia, we used the Brief Negative Symptoms Scale (BNSS) (1), a semi-structured interview conducted by trained researchers/clinicians. The BNSS is a second-generation instrument based on the NIMH-MATRICS consensus statement that is now widely recommended (2).

### Acquisition of neuroimaging data

MRI data for the main cohort were acquired at the Campus Biotech (Geneva, Switzerland) using a Siemens 3T Magnetom Prisma scanner and a standard 64 channel head coil and high-resolution resting-state images were captured with specified parameters (voxel size: 2.0 × 2.0 × 2.0 mm^3^, repetition time = 1000 ms). Full details of acquisition parameters of neuroimaging scans are available in the Supplemental Materials. MRI data for the validation cohort were acquired with the same acquisition protocol.

Images then underwent standard preprocessing involving the following steps: 1) quality assessment and realignment of functional images, 2) co-registration and segmentation using DARTEL, 3) spatial normalization to MNI space and smoothing with a 6 mm full with at half maximum Gaussian kernel, 4) exclusion of the first five scans for signal stabilization, 5) linear detrending and nuisance regression using DPARSF tissue masks (33), 6) motion correction through scrubbing of high-motion frames and 7) quality check (see Supplemental Materials for full details).

### Extraction of VTA-cerebellum CAPs and their spatiotemporal properties

Neural activity during rest is organized into time-varying, recurrent patterns of cross-regional interactions that can be captured via dynamic resting-state functional connectivity approaches. In this study, we examined the dynamic connectivity between the cerebellum and VTA through coactivation pattern (CAP) analysis (28,29), an established approach that has already contributed to the study of various psychiatric disorders (34,35) and is implemented in the publicly available *TbCAPs* toolbox (36) (Figure S1 in Supplemental Materials). In brief, timepoints with pronounced VTA activity are identified. Then, cerebellar activity at the same timepoints is retained and clustered into a set of spatial patterns, one of which is expressed at each of the retained timepoints. These patterns indicate functionally relevant networks within the cerebellum during episodes of VTA activation.

We identified the VTA using the left and right VTA masks of the Automated Anatomical Labeling (AAL) atlas (37).

For each rs-fMRI session, the BOLD signal time-series from voxels within the bilateral VTA were extracted, averaged, and standardized (Z-scored). We selected timepoints with an average value exceeding a defined threshold. Consistent with the literature, a threshold of 0.5 was employed to allow the inclusion of more timepoints and ensure robust clustering at T1, when the biggest cohort of participants was involved (38). For the smaller cohort at T2 and for the validation sample, we employed a more stringent threshold of 1 to focus only on the timepoints with largest VTA activity (36,39). Only timepoints with framewise displacement lower than 0.5 mm (scrubbing) were retained, in line with current recommendations (28).

Selected volumes underwent *K*-means clustering to extract cerebellar CAPs. Clustering was run separately on the main cohort at T1, and on the validation cohort. In both cases, the optimal number of clusters was inferred from a train-and-test approach, resulting in four CAPs in the main cohort and seven CAPs in the validation cohort (see Supplemental Materials for details). Clustering was then also run on the main cohort at T2 for four CAPs.

To quantify the longitudinal stability of CAPs, we calculated pairwise Pearson correlations between the maps obtained at T1 and at T2. Scatterplots of the correlations are provided for each group and cohort in the Supplemental Materials (Figures S4 and S5).

All CAPs were visualized using cerebellar flatmaps (40). Voxel-wise signal values were converted to t-values, using the formula:

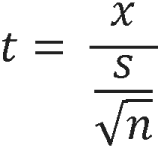

where *x* is the average BOLD value for the voxel at hand across the frames in which the CAP of interest is expressed, *s* is the standard deviation, and *n* is the number of frames. These maps effectively quantify the extent and stability of voxel activity within each dynamic state.

Two complementary metrics reflective of CAP temporal dynamics were calculated as described in (36) (Figure S1): *persistence*, or the average time, in seconds, a CAP was consecutively expressed upon occurrence, and *emergence*, denoting the frequency, irrespective of duration, at which a specific CAP occurred after VTA under-threshold activity.

### Exploration of the relation of CAP metrics with negative symptoms

Statistical analyses included non-parametric testing with rank tests for median comparisons to contrast CAP metrics across groups, and Spearman’s correlation to examine the associations of these metrics with psychopathology. Non-parametric testing allows for analyses without assumptions about data distribution. To evaluate the statistical significance of partial correlations given the relatively small sample size, non-parametric permutation testing with 5000 iterations was also employed (see Supplemental Materials for more details). These analyses were conducted in Python 3.9 using *statsmodels* (version 0.14.1). Throughout this work, the False Discovery Rate (FDR) was controlled using the Benjamini-Hochberg procedure to adjust p-values for multiple comparisons. All significant results survive FDR correction at a significant level of p<0.05 unless reported otherwise. To account for movement in the scanner and medication intake, unless specified otherwise, framewise displacement and risperidone equivalent dose were included as covariates in group comparisons of CAP metrics and partial correlation analyses with psychopathology through ANCOVA as implemented in Python 3.9 using the *statsmodels* and *pingouin* libraries (version 0.14.1).

For the main cohort at T1, the HC and SZ groups were contrasted, for each CAP, in terms of emergence and persistence. CAPs for which a significant group difference was found were further examined in terms of associations of the metrics with the severity of negative symptoms (apathy and diminished expression) as measured by the BNSS. A similar process was then performed at T2, and on the validation dataset, on the CAPs matching those of interest brought about at T1. Note that in the validation dataset, we examined the specific hypothesis derived from the main cohort through one-tailed tests.

The code for the above analyses can be found at: https://drive.google.com/drive/folders/1roAKf33fQ1S_q0uajCnlcPQvlV8-Th6t?usp=drive_link. All figures in this manuscript are generated in Biorender (https://BioRender.com).

## RESULTS

### Demographic, neuroimaging, and clinical data in the main cohort

A summary of the main cohort characteristics from T1 to T3 can be found in Table 1. In this cohort, the patient population was matched for age and sex to healthy controls and included chronic and clinically stable patients with low levels of psychosis, depression and extrapyramidal syndrome. Negative symptoms in the patients’ cohort were enduring from T1 to T3 and not associated with psychosis, medication, cognitive dysfunction, or extrapyramidal effects, suggesting that these symptoms were unconfounded by other clinical factors (see Supplemental Materials). This allows us to obtain a mechanistic understanding of the biological mechanisms driving negative symptoms.

**Table 1.**
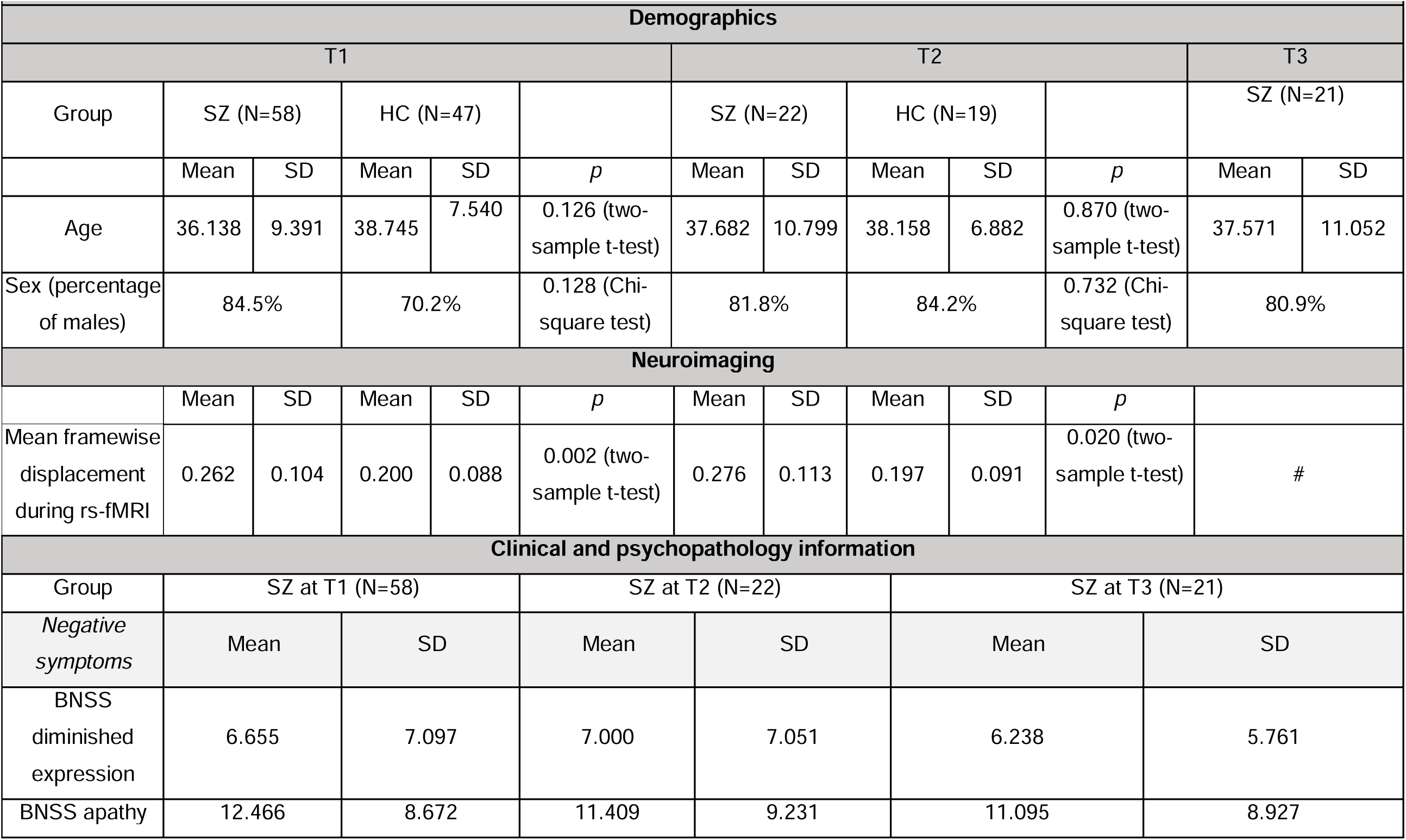
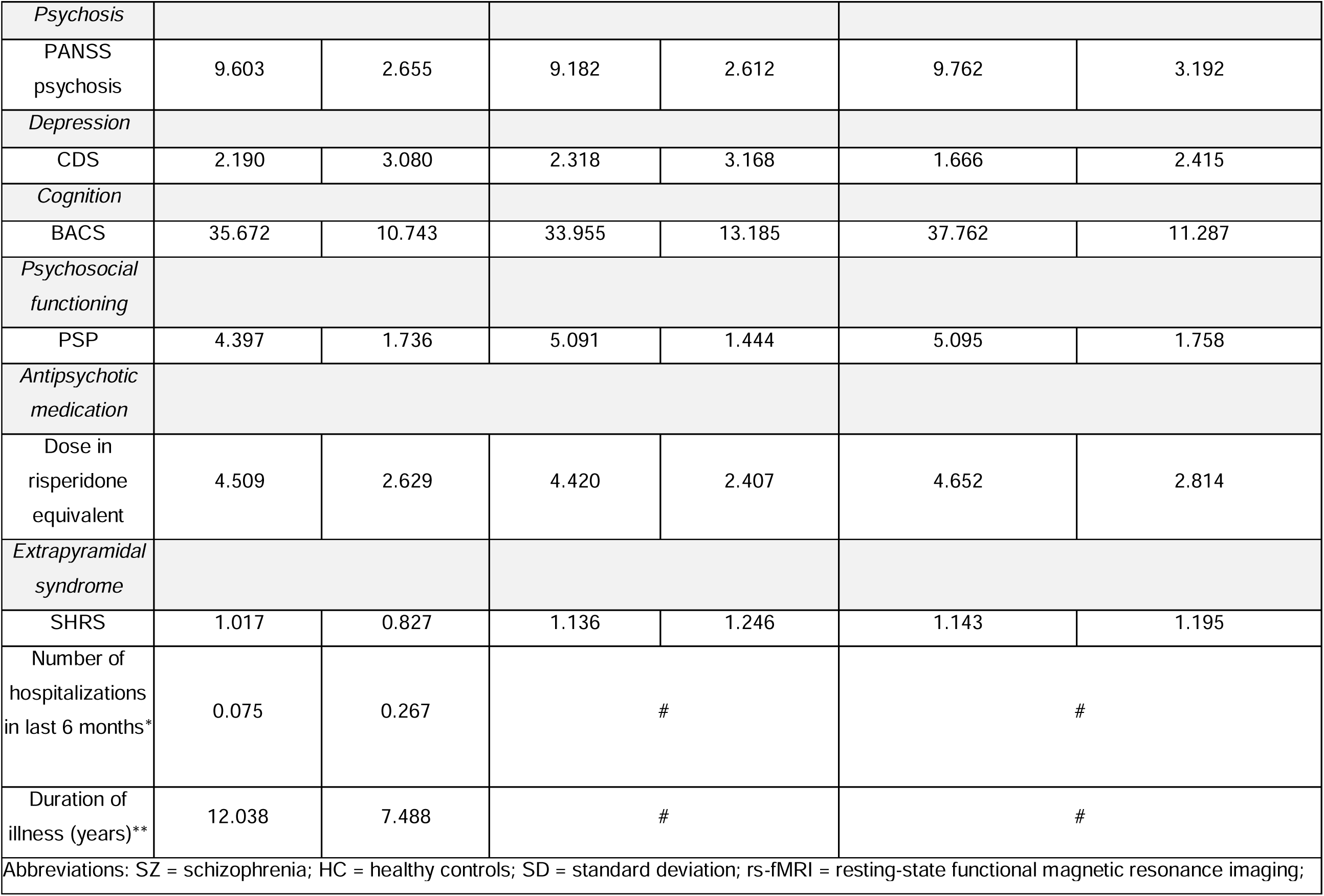
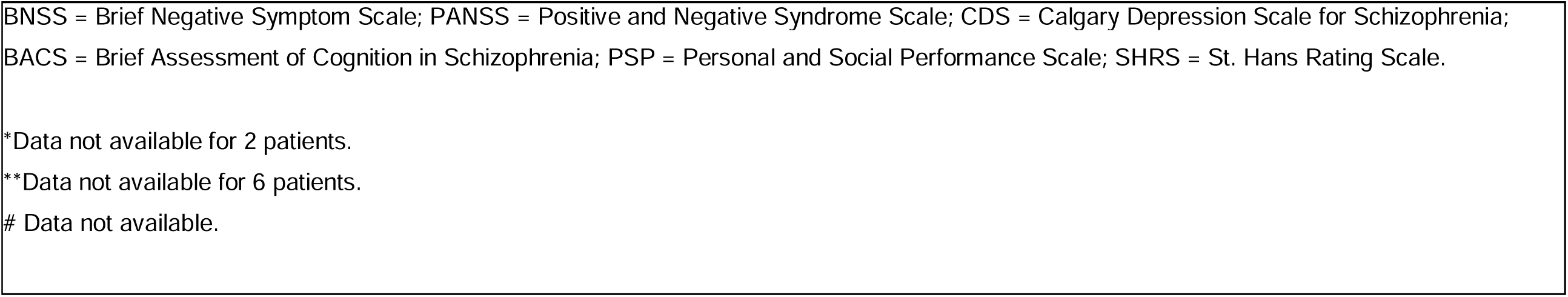
Demographic and psychopathology characteristics of study participants (main cohort)

|  |  |  |  |  |  |  |  |  |  |  |  |  |
| --- | --- | --- | --- | --- | --- | --- | --- | --- | --- | --- | --- | --- |
| Demographics |  |  |  |  |  |  |  |  |  |  |  |  |
| T1 |  |  |  |  |  | T2 |  |  |  |  | T3 |  |
| Group | SZ (N=58) |  | HC (N=47) |  |  | SZ (N=22) |  | HC (N=19) |  |  | SZ (N=21) |  |
|  | Mean | SD | Mean | SD | *p* | Mean | SD | Mean | SD | *p* | Mean | SD |
| Age | 36.138 | 9.391 | 38.745 | 7.540 | 0.126 (two-sample t-test) | 37.682 | 10.799 | 38.158 | 6.882 | 0.870 (two-sample t-test) | 37.571 | 11.052 |
| Sex (percentage of males) | 84.5% |  | 70.2% |  | 0.128 (Chi-square test) | 81.8% |  | 84.2% |  | 0.732 (Chi-square test) | 80.9% |  |
| Neuroimaging |  |  |  |  |  |  |  |  |  |  |  |  |
|  | Mean | SD | Mean | SD | *p* | Mean | SD | Mean | SD | *p* |  |  |
| Mean framewise displacement during rs-fMRI | 0.262 | 0.104 | 0.200 | 0.088 | 0.002 (two-sample t-test) | 0.276 | 0.113 | 0.197 | 0.091 | 0.020 (two-sample t-test) | # |  |
| Clinical and psychopathology information |  |  |  |  |  |  |  |  |  |  |  |  |
| Group | SZ at T1 (N=58) |  |  |  | SZ at T2 (N=22) |  |  |  | SZ at T3 (N=21) |  |  |  |
| *Negative symptoms* | Mean |  | SD |  | Mean |  | SD |  | Mean |  | SD |  |
| BNSS diminished expression | 6.655 |  | 7.097 |  | 7.000 |  | 7.051 |  | 6.238 |  | 5.761 |  |
| BNSS apathy | 12.466 |  | 8.672 |  | 11.409 |  | 9.231 |  | 11.095 |  | 8.927 |  |

|  |  |  |  |  |  |  |
| --- | --- | --- | --- | --- | --- | --- |
| *Psychosis* |  |  |  |  |  |  |
| PANSS psychosis | 9.603 | 2.655 | 9.182 | 2.612 | 9.762 | 3.192 |
| *Depression* |  |  |  |  |  |  |
| CDS | 2.190 | 3.080 | 2.318 | 3.168 | 1.666 | 2.415 |
| *Cognition* |  |  |  |  |  |  |
| BACS | 35.672 | 10.743 | 33.955 | 13.185 | 37.762 | 11.287 |
| *Psychosocial functioning* |  |  |  |  |  |  |
| PSP | 4.397 | 1.736 | 5.091 | 1.444 | 5.095 | 1.758 |
| *Antipsychotic medication* |  |  |  |  |  |  |
| Dose in risperidone equivalent | 4.509 | 2.629 | 4.420 | 2.407 | 4.652 | 2.814 |
| *Extrapyramidal syndrome* |  |  |  |  |  |  |
| SHRS | 1.017 | 0.827 | 1.136 | 1.246 | 1.143 | 1.195 |
| Number of hospitalizations in last 6 months* | 0.075 | 0.267 | # |  | # |  |
| Duration of illness (years)** | 12.038 | 7.488 | # |  | # |  |
| Abbreviations: SZ = schizophrenia; HC = healthy controls; SD = standard deviation; rs-fMRI = resting-state functional magnetic resonance imaging; |  |  |  |  |  |  |

BNSS = Brief Negative Symptom Scale; PANSS = Positive and Negative Syndrome Scale; CDS = Calgary Depression Scale for Schizophrenia; BACS = Brief Assessment of Cognition in Schizophrenia; PSP = Personal and Social Performance Scale; SHRS = St. Hans Rating Scale.
\*Data not available for 2 patients.
\*\*Data not available for 6 patients.

### Investigation of cerebellar temporal coupling with the VTA at T1

Our neuroimaging analysis uncovered four spatially distinct cerebellum–VTA CAPs (Figure 1A; refer to the Supplemental Materials for the selection of the optimal number of clusters).

**Figure 1.**
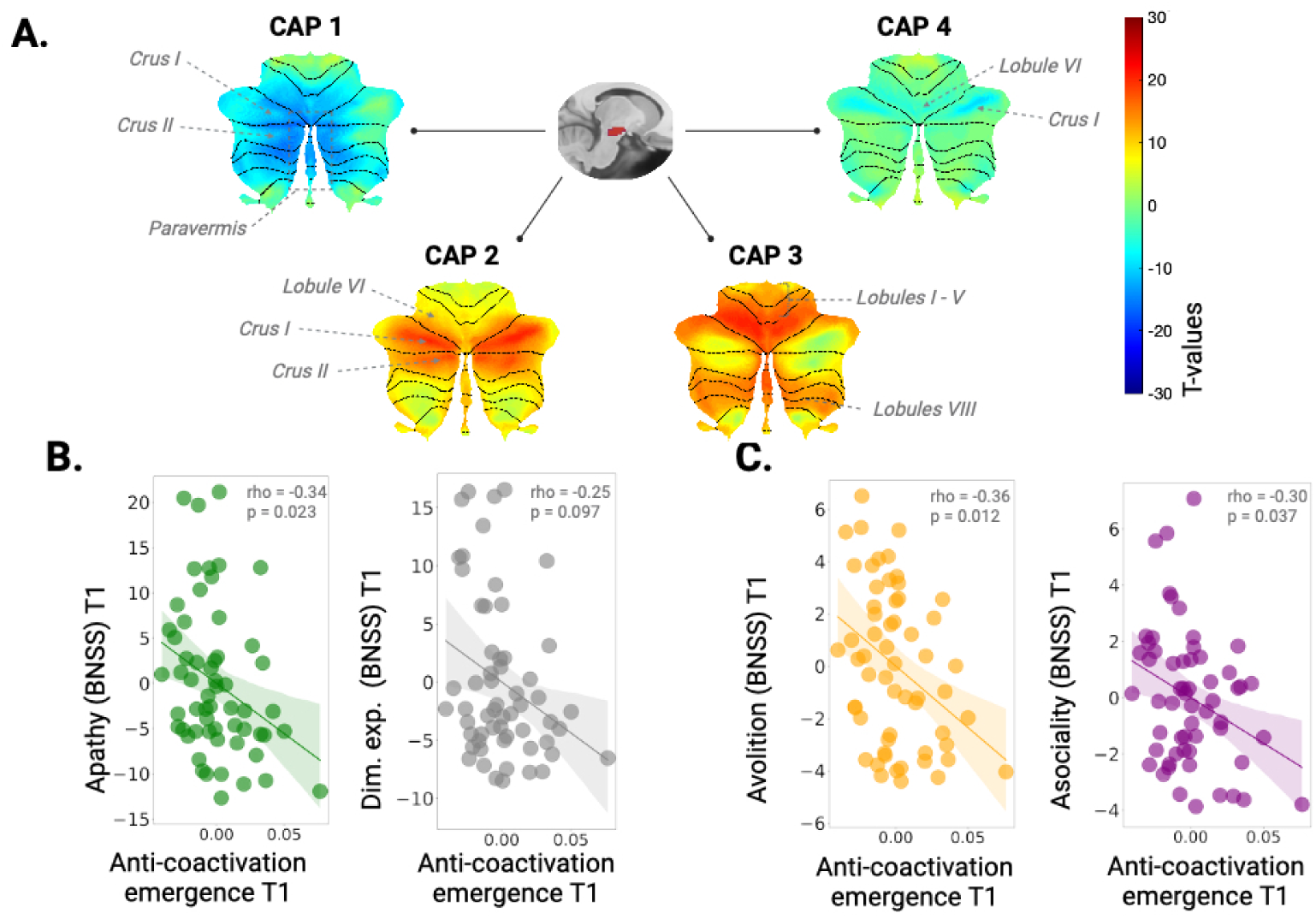
Spatial patterns of coactivation between the cerebellum and the VTA in patients (SZ) and controls (HC) and their relation to negative symptoms at T1. A. Distinct spatial maps of cerebellar coactivation with the VTA. B. Reduced anti-coactivation pattern emergence (CAP1 entry frequency) is associated with greater severity of apathy (in green) but not diminished expression (in grey). C. Reduced anti-coactivation pattern emergence (CAP1 entry frequency) is associated with greater severity of avolition (in yellow) and of asociality (in purple). *Note*. *Negative symptoms quantified by the Brief Negative Symptom Scale (BNSS). Greater scores indicate more severe symptoms. Dim. exp. = diminished expression. The colorbar represents T-values. Correlations are corrected for movement (mean framewise displacement) and antipsychotic medication (risperidone equivalent). Shaded areas on correlation plots B. and C. represent the 95% confidence interval*.

CAP1 was identified as a cerebellar anti-coactivation pattern with the VTA and was strongest at bilateral paravermal regions of Crus I and II. CAP2 showed coactivation with the VTA within lobule VI extending to Crus I and II. CAP3 encompassed mainly cerebellar signal in the anterior (lobules I-V) and posterior (lobules VIIIA and VIIIB) lobes. Finally, CAP4 showed anti-coactivation of the VTA predominantly with lateral Crus I and lobule VI.

#### At T1, CAP1 persistence is reduced in schizophrenia and its emergence relates to apathy but not diminished expression

We compared each CAP’s persistence (average duration in seconds) and emergence (number of times a specific pattern emerges after VTA under-threshold activity) between patients with schizophrenia and HC and, found a significantly shorter CAP1 duration in patients (F(1,103) = 16.13, p = 0.0009, without covariates). Correction for antipsychotic dosage in patients (quantified by risperidone equivalents) and mean framewise displacement did not affect this result (F(1,101) = 5.609, p = 0.02). We found no significant differences in the persistence of the remaining three CAPs across the two diagnostic groups. Further, we found no significant differences in CAP emergence between the groups. Therefore, we focused on the clinical relevance of CAP1 for patients, by assessing the association between its temporal features (emergence and persistence) and negative symptoms (apathy and diminished expression as quantified by the BNSS subscales).

We found that CAP1 persistence was not associated with apathy or diminished expression. CAP1 emergence was significantly negatively correlated with the severity of apathy (*rho* =−0.31, p = 0.035; Figure 1B, left panel, without covariates), while the association with the severity of diminished expression was not significant (*rho* =−0.25, p = 0.099; Figure 1B, right panel, without covariates). Correction for antipsychotic dosage (quantified by risperidone equivalents) and mean framewise displacement yielded similar results (apathy: *rho* =−0.34, p = 0.023; diminished expression: *rho* =−0.25, p = 0.097).

To gain a more detailed understanding of the relationship with apathy, we examined the individual apathy symptoms (i.e., avolition, asociality, and anhedonia) and observed that CAP1 emergence was associated with avolition (*rho* =−0.36, p = 0.012; Figure 1C, left panel) and asociality (*rho* =−0.3, p = 0.037; Figure 1C, right panel), but not with anhedonia (*rho* =−0.24, p = 0.102). Non-parametric permutation testing also confirmed these results (apathy: *rho* =−0.34, p = 0.012; avolition: *rho* =−0.36, p = 0.007; and asociality: *rho* =−0.30, p = 0.024) (see Figure S9 in the Supplemental Materials).

### Investigation of cerebellar temporal coupling with the VTA at T2

#### Reproducible anti-coactivation pattern of the cerebellum with VTA in two distinct recordings

Like at T1, we identified four distinct cerebellum-VTA CAPs at T2 (Figure 2A). Comparison of spatial similarity at T1 and T2 using pairwise spatial correlation revealed an almost identical spatial equivalent of the cerebellar anti-coactivation (*r* = 0.98, p < 0.001) at T2, in favor of a reproducible functional signature (Figure 2B; see also Figures S4 and S5 in Supplemental Materials). For simplicity, we also refer to this almost identical pattern at T2 as CAP1 in what follows.

**Figure 2.**
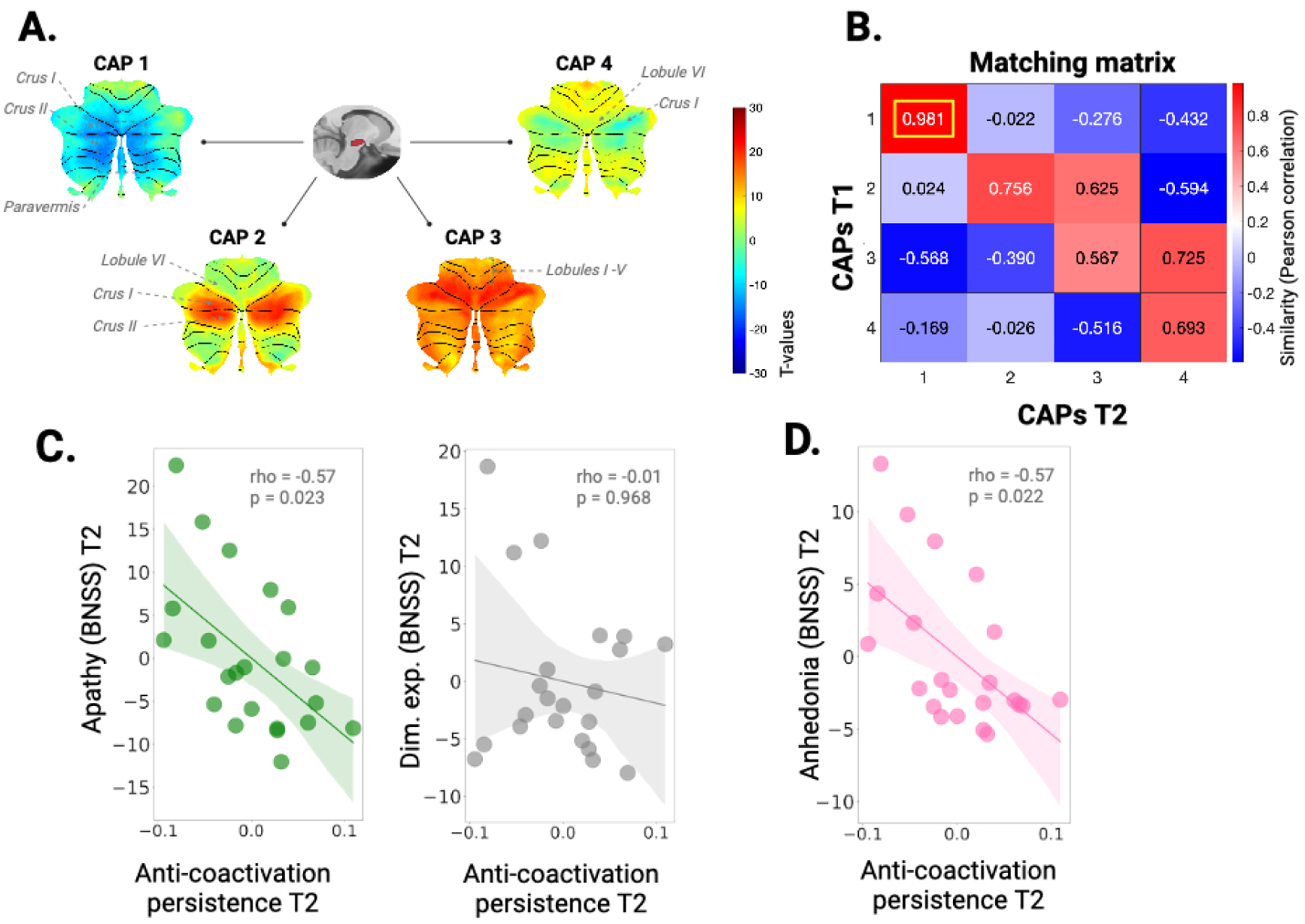
Spatial patterns of coactivation between the cerebellum and the VTA in patients (SZ) and controls (HC) and their relation to negative symptoms at T2. A. Distinct spatial maps of cerebellar coactivation with the VTA. B. Correlation matrix between patterns of coactivation at T1 and at T2. C. Reduced anti-coactivation persistence (CAP1 duration in seconds) is associated with greater severity of apathy (in green) but not diminished expression (in grey). D. Reduced anti-coactivation persistence is associated with greater severity of anhedonia (in pink). *Note*. *Negative symptoms are quantified by the Brief Negative Symptom Scale (BNSS). Greater scores indicate more severe symptoms. Dim. exp. = diminished expression. The colorbar in panel A represents T-values, while that in panel B depicts similarity (Pearson correlation). Correlations are corrected for movement (mean framewise displacement) and antipsychotic medication (risperidone equivalent). Shaded areas on correlation plots C. and D. represent the 95% confidence interval*.

#### Anti-coactivation pattern persistence is reduced in schizophrenia and relates to apathy at T2

We examined the anti-coactivation pattern persistence and emergence between patients with schizophrenia and HC, and like at T1, found a significantly shorter duration in patients at T2 (F(1,29) = 6.97, p = 0.026). Persistence was also significantly negatively correlated with the severity of apathy (*rho* =−0.57, p = 0.023; Figure 2C, left panel) but not of diminished expression (*rho* =−0.01, p = 0.968; Figure 2C, right panel). By contrast, CAP1 emergence did not relate to negative symptom severity.

Again, we sought to obtain a more granular understanding of the relationship with apathy at T2, and examined the association with the individual items: anhedonia, asociality and avolition. We observed that CAP1 persistence was associated with anhedonia (*rho* =−0.57, p = 0.022), but neither with asociality nor with avolition (*rho* =−0.4, p = 0.167, *rho* =−0.3, p = 0.308, respectively). In sum, these findings suggest a distinct mapping of apathy subdomains to specific anti-coactivation metrics, with persistence putatively linked to anhedonia (as seen at T2) and emergence potentially associated with asociality or avolition (as observed at T1). We replicated the results using non-parametric permutations testing (apathy: *rho* =−0.57, p = 0.008; anhedonia: *rho* =−0.57, p = 0.009) (see Figure S10 in the Supplemental Materials).

### Anti-coactivation pattern metrics at T2 are associated with apathy at T3

Finally, we asked whether CAP1 metrics at T1 or at T2 could be linked to symptoms at T3. We found that CAP1 metrics at T1 were not associated with apathy or diminished expression at T2 or T3. By contrast, the persistence of the anti-coactivation pattern at T2 correlated negatively with the severity of apathy at T3 (*rho* =−0.48, p = 0.049; Figure 3A, left panel), but not with that of diminished expression (*rho* =−0.2, p = 0.412; Figure 3A, right panel).

**Figure 3.**
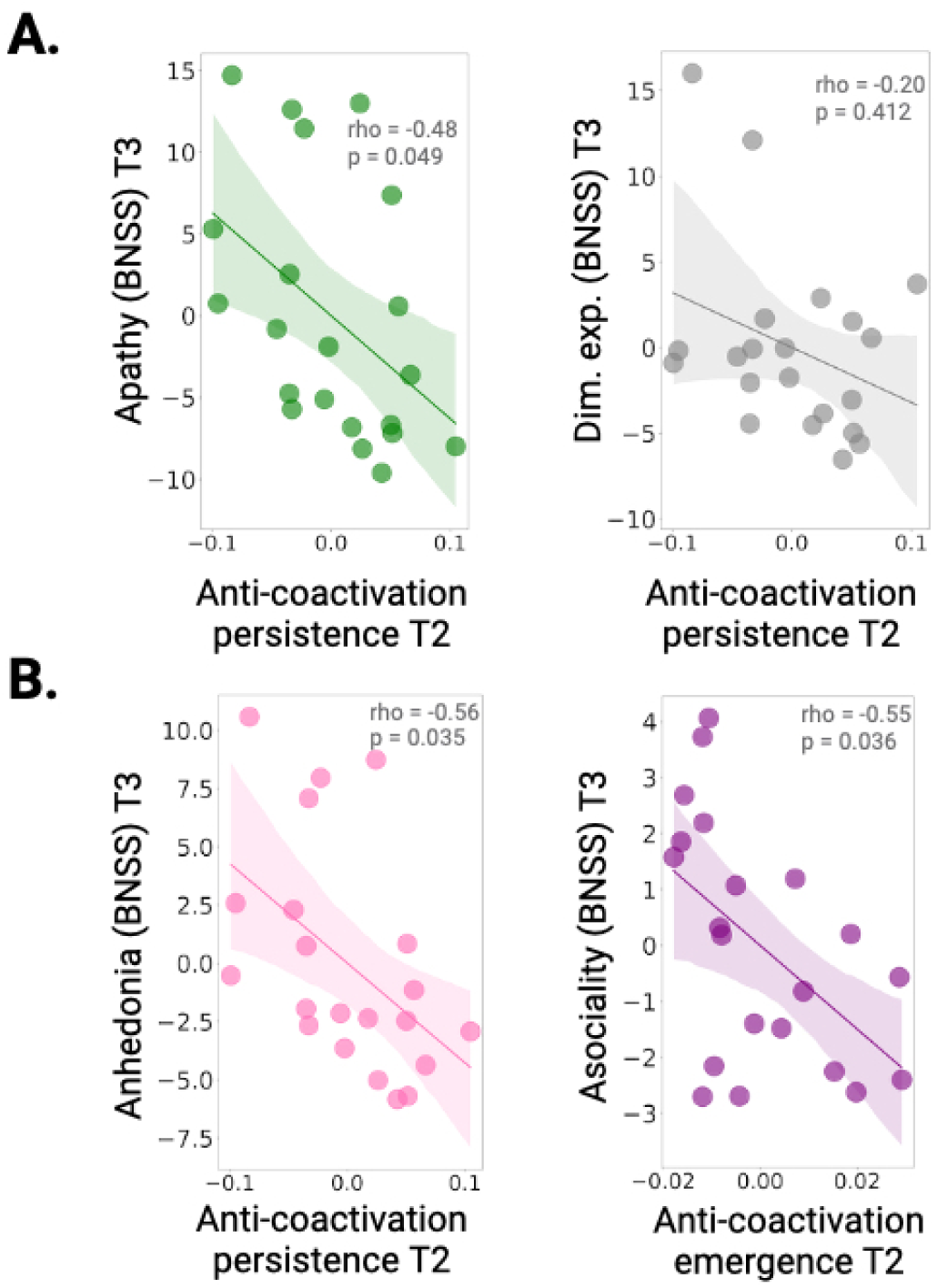
Relation between anti-coactivation metrics at T2 and negative symptoms at T3. A. Reduced anti-coactivation persistence (CAP1 duration in seconds) is associated with greater severity of apathy (in green) but not diminished expression (in grey). B. Reduced anti-coactivation persistence is associated with greater severity of anhedonia (in pink), while reduced anti-coactivation emergence (CAP1 entry frequency) is associated with greater severity of asociality (in purple). *Note. Negative symptoms are quantified by the Brief Negative Symptom Scale (BNSS). Greater scores indicate more severe symptoms. Dim. exp. = diminished expression. Correlations are corrected for movement (mean framewise displacement) and antipsychotic medication (risperidone equivalent). Shaded areas on correlation plots represent the 95% confidence interval*.

A more detailed examination uncovered that CAP1 *persistence* related to anhedonia (*rho* =−0.56, p = 0.035; Figure 3B, left panel) while CAP1 *emergence* related to asociality (*rho* =−0.55, p = 0.036; Figure 3B, right panel), replicating the previous findings of a differential relationship of anti-coactivation metrics with distinct negative symptoms observed at T1 and T2. Non-parametric permutations testing confirmed these results (apathy: *rho* =−0.48, p = 0.042; anhedonia: *rho* =−0.56, p = 0.014; asociality: *rho* =−0.55, p = 0.013)(see Figure S11 in the Supplemental Materials).

### Validation dataset analyses

A summary of the validation cohort characteristics across timepoints can be found in Table 2.

**Table 2.**
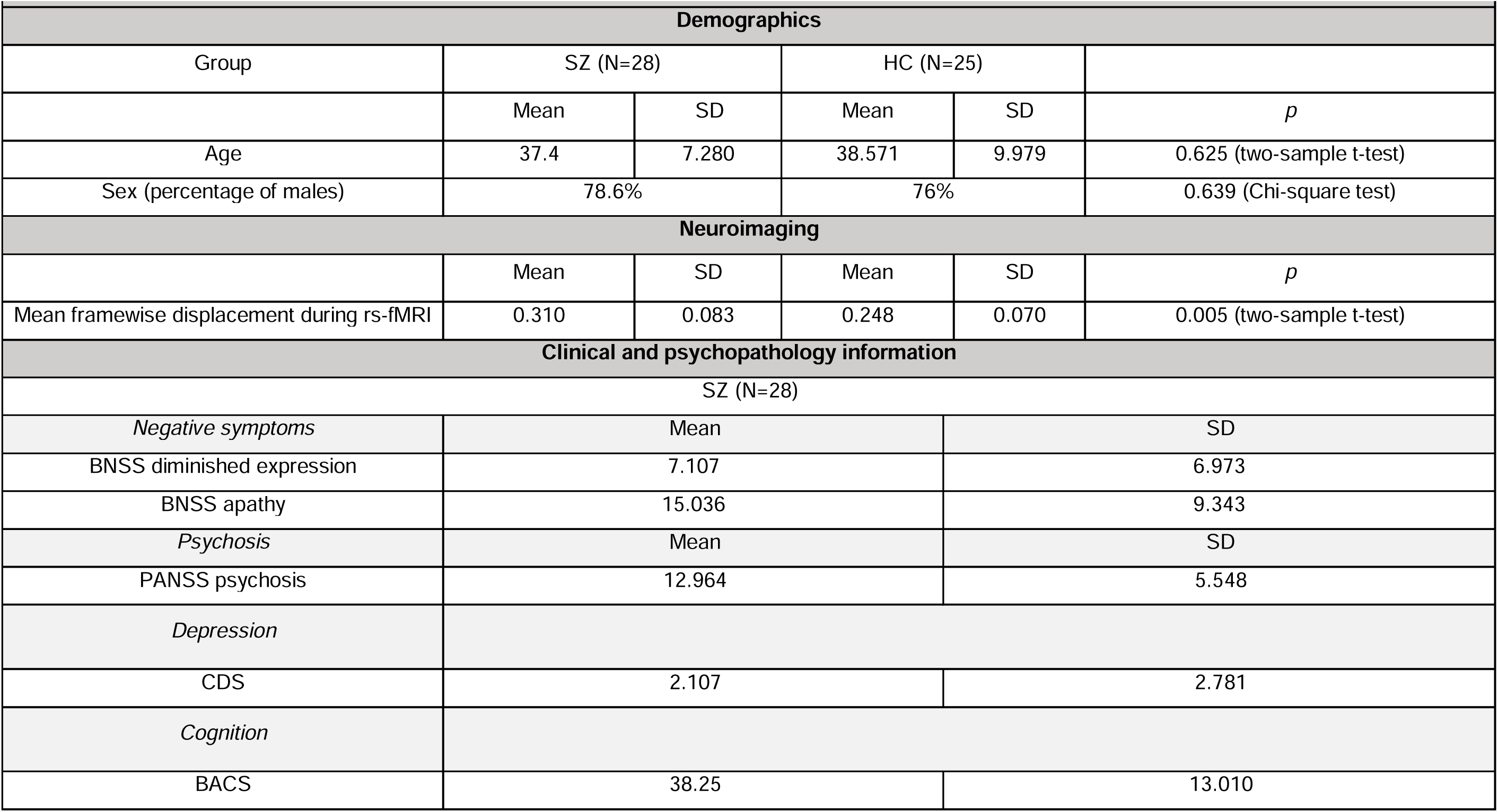
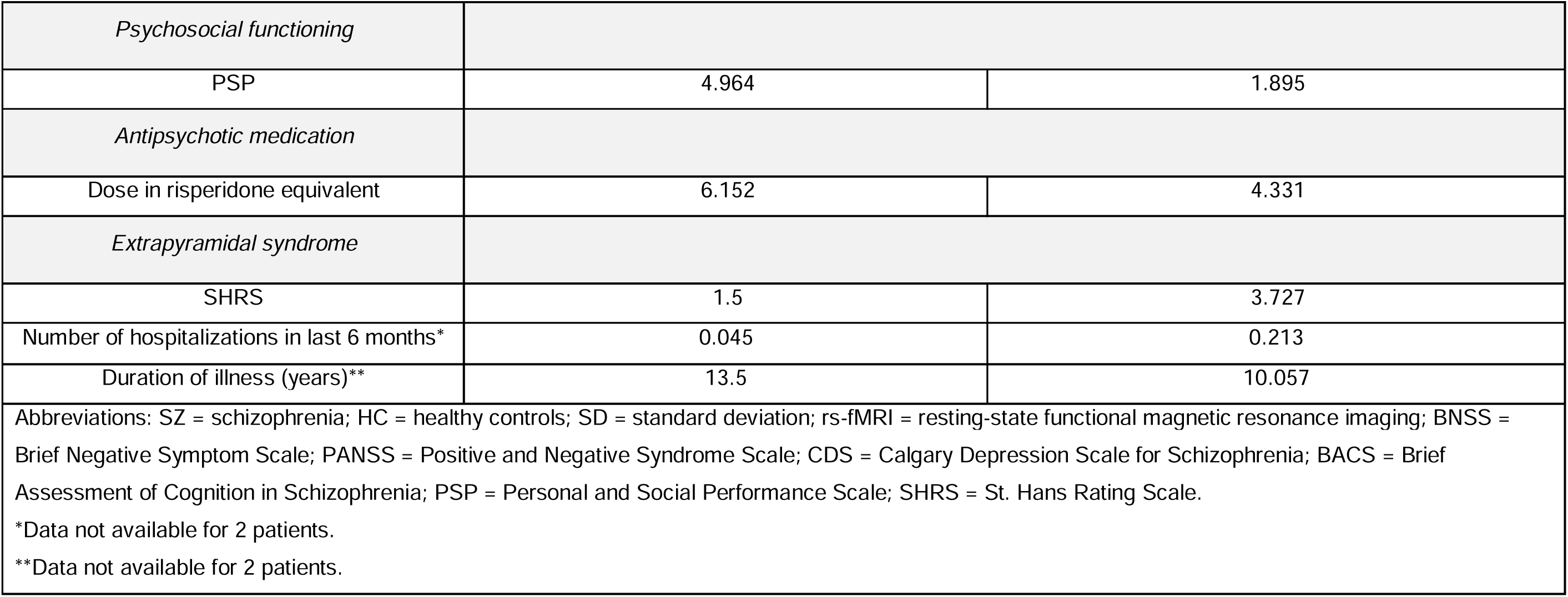
Demographic and psychopathology characteristics of study participants (validation cohort)

|  |  |  |  |  |  |
| --- | --- | --- | --- | --- | --- |
| Demographics |  |  |  |  |  |
| Group | SZ (N=28) |  | HC (N=25) |  |  |
|  | Mean | SD | Mean | SD | *p* |
| Age | 37.4 | 7.280 | 38.571 | 9.979 | 0.625 (two-sample t-test) |
| Sex (percentage of males) | 78.6% |  | 76% |  | 0.639 (Chi-square test) |
| Neuroimaging |  |  |  |  |  |
|  | Mean | SD | Mean | SD | *p* |
| Mean framewise displacement during rs-fMRI | 0.310 | 0.083 | 0.248 | 0.070 | 0.005 (two-sample t-test) |
| Clinical and psychopathology information |  |  |  |  |  |
| SZ (N=28) |  |  |  |  |  |
| *Negative symptoms* | Mean |  |  | SD |  |
| BNSS diminished expression | 7.107 |  |  | 6.973 |  |
| BNSS apathy | 15.036 |  |  | 9.343 |  |
| *Psychosis* | Mean |  |  | SD |  |
| PANSS psychosis | 12.964 |  |  | 5.548 |  |
| *Depression* |  |  |  |  |  |
| CDS | 2.107 |  |  | 2.781 |  |
| *Cognition* |  |  |  |  |  |
| BACS | 38.25 |  |  | 13.010 |  |

|  |  |  |
| --- | --- | --- |
| *Psychosocial functioning* |  |  |
| PSP | 4.964 | 1.895 |
| *Antipsychotic medication* |  |  |
| Dose in risperidone equivalent | 6.152 | 4.331 |
| *Extrapyramidal syndrome* |  |  |
| SHRS | 1.5 | 3.727 |
| Number of hospitalizations in last 6 months* | 0.045 | 0.213 |
| Duration of illness (years)** | 13.5 | 10.057 |
| Abbreviations: SZ = schizophrenia; HC = healthy controls; SD = standard deviation; rs-fMRI = resting-state functional magnetic resonance imaging; BNSS = Brief Negative Symptom Scale; PANSS = Positive and Negative Syndrome Scale; CDS = Calgary Depression Scale for Schizophrenia; BACS = Brief Assessment of Cognition in Schizophrenia; PSP = Personal and Social Performance Scale; SHRS = St. Hans Rating Scale.<br>*Data not available for 2 patients.<br>**Data not available for 2 patients. |  |  |

The validation cohort’s population was matched for age and sex to healthy controls and included chronic and clinically stable patients with similar levels of negative symptoms to the main cohort (see Table S4 in Supplemental Materials). Both cohorts were also similar for other clinical parameters like medication, cognitive dysfunction, or extrapyramidal effects. However, patients in the validation cohort had relatively higher psychosis scores compared to patients in the main cohort (see Tables S4 and S5 in Supplemental Materials for uncorrected results).

Investigation of cerebellar temporal coupling with the VTA in the validation cohort revealed seven spatially distinct cerebellum–VTA CAPs (Figure 4A; see Supplemental Materials for the selection of the optimal number of clusters). A prominent anti-coactivation pattern was again observed in the medial paravermal regions of Crus I and II. Pairwise spatial correlation analysis comparing the similarity of this anti-coactivation pattern with that from the main cohort at T1 confirmed spatial consistency (*r* = 0.93, p < 0.001; Figure 4B), suggesting a reproducible functional signature.

**Figure 4.**
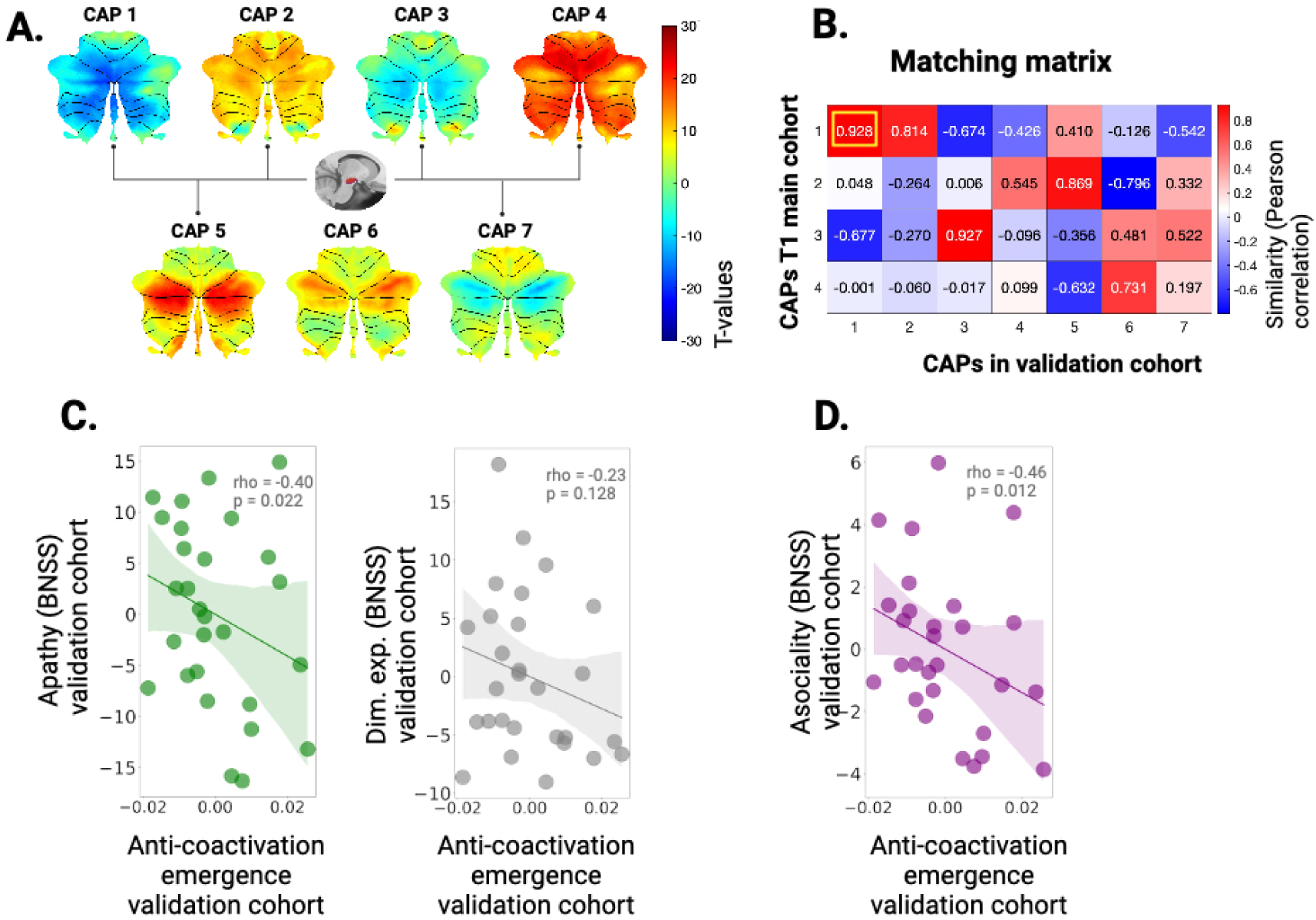
Validation cohort analysis: relation between anti-coactivation emergence and negative symptoms. A. Distinct spatial maps of cerebellar coactivation with the VTA in the validation cohort. B. Correlation matrix between coactivation patterns at T1 in the main cohort and coactivation patterns at T1 in the validation cohort. C. Reduced anti-coactivation emergence (CAP1 entry frequency) is associated with greater severity of apathy (in green). D. Reduced anti-coactivation emergence (CAP1 entry frequency) is associated with greater severity of asociality (in purple). *Note. Negative symptoms are quantified by the Brief Negative Symptom Scale (BNSS). Greater scores indicate more severe symptoms. Dim. exp. = diminished expression. The colorbar in panel A represents T-values, while that in panel B depicts similarity (Pearson correlation). Correlations are corrected for movement (mean framewise displacement) and antipsychotic medication (risperidone equivalent). Shaded areas on correlation plots C. and D. represent the 95% confidence interval*

We examined the anti-coactivation pattern persistence and emergence between patients with schizophrenia and HC. We found no significant between-group differences in the metrics of this pattern. However, the anti-coactivation pattern emergence was significantly negatively correlated with the severity of apathy (*rho* =−0.4, p= 0.022; Figure 4C, left panel), but not of diminished expression (*rho* =−0.23, p= 0.128; Figure 4C, right panel), replicating our findings of the main cohort at T1. However, no associations with apathy were found for the pattern’s persistence.

In a more granular examination, we found that the anti-coactivation pattern emergence in the validation dataset related to asociality (rho =−0.46, p= 0.012), replicating our findings from the main cohort. Of note, the relationship of the anti-coactivation pattern emergence with avolition was also close to significance (rho =−0.32, p= 0.066). We replicated these results using non-parametric permutations testing (apathy: *rho* =−0.40, p = 0.043; asociality: *rho* =−0.46, p = 0.017) (see Figure S12 in the Supplemental Materials).

## DISCUSSION

We uncovered a longitudinally stable spatial functional signature of anti-coactivation involving the cerebellar vermis and paravermis regions within Crus I – Crus II and the VTA in schizophrenia. This pattern was replicated into two distinct recordings (i.e., at baseline and at 3-month follow-up; Figure 2B) as well as in an independent sample with very high concordance (Figure 4B). Patients with scarcer emergence or reduced persistence of anti-coactivation had more severe apathy, suggesting that this pattern may be important for motivated behavior. Further, we observed a facetted relationship between features of the anti-coactivation and components of apathy: emergence appeared to more related to deficits of *wanting* – the drive to pursue a reward or goal, as indicated by its association with avolition and asociality, both reflecting reductions in goal-directed behavior or social motivational drive. In contrast, its persistence showed a stronger link to deficits of *liking* – the pleasure or positive emotional response derived from achieving or consuming a reward, specifically through anhedonia, reflecting diminished capacity for pleasure or positive emotional response. This distinction suggests that motivational and hedonic aspects may engage uniquely with anti-coactivation dynamics and evolve differently over time in ways that remains to be fully elucidated. Thus, anti-coactivation may serve as a biomarker for these distinct facets of apathy ultimately informing targeted interventions for specific components of apathy in schizophrenia that should be the focus of future research.

Our findings concord with evidence of a dysfunctional cerebellar ‘inhibitory tone’ over deeper structures (16,21) that could contribute to the excitatory/inhibitory (E/I) imbalance observed in schizophrenia (41). The balance between neuronal excitation and inhibition is crucial for optimizing the signal-to-noise ratio within neuronal networks (42). Dysregulation of cerebellar neurons projecting to the midbrain would disrupt the E/I and the signal-to-noise ratios, thereby diminishing the brain’s ability to distinguish between rewarding and non-rewarding stimuli and thus leading to apathy (41). Given its anatomical structure with excitatory (*i.e.*, granule cells) and inhibitory neurons (*i.e.*, Purkinje cells), the cerebellum is a central hub for regulating both inhibition and excitation. The cerebellar Purkinje cells that coordinate the glutamatergic cerebellar output, have very high metabolic energy demands which makes them susceptible to damage (43) and thus prone to disturbing the E/I balance upon exposure to genetic and environmental stressors. Post-mortem studies of individuals with autism – a disorder also presenting with amotivated behavior and clinical overlap with schizophrenia – corroborated a reduced number of Purkinje cells, as well as alterations in cerebellar volume and connectivity (44). In sum, the cerebellum’s critical role in gatekeeping cortical neuronal excitation and inhibition, coupled with its vulnerability to genetic and environmental stressors, points to its key role in the pathophysiology of amotivated behavior across schizophrenia and other developmental disorders.

In our study, we observed that features of the anti-coactivation pattern expressed in the *medial* (i.e., CAP1) but not the lateral cerebellum (i.e., CAP4) related to apathy over time (T2, T3). The implication of the medial cerebellum in the paravermis area provides further support for the Cognitive Dysmetria theory that postulated a key role of these regions in schizophrenia (45). The vermis and paravermis regions of Crus I and II, unlike their lateral counterparts, have unique molecular properties that may confer resilience to stress, enabling compensatory processes that require time in line with our findings (Figure 3). Specifically, Purkinje cells in these posterior cerebellar areas express zebrin-II, a highly conserved protein across vertebrates that provides resistance to injury (46), unlike the motor cerebellum, which is negative for zebrin-II and more susceptible to stress. This would imply that developmental stress impacts the motor cerebellum before non-motor areas. In this context, schizophrenia’s clinical presentation could be viewed through cerebellar zebrin patterns, with initial “soft” neurological dysfunctions such as ataxia and dysmetria relying on motor areas (47) preceding negative and cognitive symptoms relying on the posterior cerebellum. Investigating the molecular mechanisms in these regions could enhance our understanding of dysfunctional cerebellar circuits in schizophrenia and should be the focus of future efforts (4,17).

Our study not only sheds light on the cerebellum regulatory role in the pathophysiology of apathy in schizophrenia, but also opens new avenues for therapeutic intervention. The specific cerebellar–VTA connectivity patterns that we identified offer promising targets for anti-coactivation-guided individualized cerebellar transcranial magnetic stimulation (TMS), aiming to restore functional neural circuits and alleviate negative symptoms in schizophrenia.

Our study has strengths and limitations. We identify for the first time and validate a longitudinally replicable anti-coactivation pattern of cerebellum–VTA connectivity in humans relating to negative symptoms in schizophrenia, bridging a critical gap on how the cerebellum regulates the reward circuitry over time. We employed an established method of dynamic functional connectivity (28,29,48–50), with strict quality control measures in place to ensure the reliability of our findings. We used a scale specifically designed to measure negative symptoms excluding items related to non-negative symptom psychopathology domains included in older scales (1,16) in line with the most recent guidelines (1). Following these guidelines, we utilized a clinical cohort with prominent negative symptoms and minimal confounding from psychosis, medication, or extrapyramidal effects. This approach enhances the validity of the mechanism-informed understanding of negative symptoms that we aimed to achieve in our work.

Regarding limitations, given that recruitment occurred during the COVID-19 pandemic, we encountered significant challenges for organizing and recruiting participants, leading to participant attrition between time points. Another limitation is the presence of predominantly male individuals, consistent with the literature and the gender repartition of schizophrenia in the population and, in particular, of negative symptoms in men (51). Future studies should examine sex differences in the expression of the patterns reported here. Finally, given our use of resting-state fMRI data, our measure of cerebellar activity is indirect. However, the blood oxygenation level-dependent signal has been linked to excitatory function (32), and therefore the observed anti-coactivation likely indicates either increased inhibition or reduced excitatory activity. To reflect this nuance, in this research we have employed the term “anti-coactivation” to remain agnostic about the exact underlying mechanism. However, future studies should aim to directly examine the specific neuronal activity and mechanisms underlying these patterns to provide a clearer understanding. In humans, this could be achieved through advanced imaging techniques like magnetoencephalography or invasive electrophysiological recordings in specific neurosurgical contexts, along with non-invasive neuromodulation techniques like TMS.

In conclusion, we identified that a longitudinally stable anti-coactivation pattern of the cerebellum with the VTA relates to apathy in schizophrenia. These results open new avenues for therapeutic interventions through resting-state anti-coactivation-targeted cerebellar neuromodulation for more effective and personalized treatments.

## Data Availability

Data will be available upon reasonable request to the authors.

## Funding

Leenaards Foundation grant awarded to IB. PRD funds (22-2020-I) awarded to IB. Swiss National Science Foundation Grant No 169783 awarded to SK.

## Contributions

Farnaz Delavari: Investigation, Methodology, Formal analysis, Writing original draft, Visualization. Jade Awada: Investigation, Formal analysis, Writing original draft, Visualization. Dimitri Van De Ville: Methodology, Review and editing of manuscript. Thomas A. Bolton: Methodology, Review and editing of manuscript. Mariia Kaliuzhna: Data acquisition. Fabien Carruzzo: Data acquisition. Noemie Kuenzi: Data acquisition. Teresa Marie Katthagen; Data Acquisition; Tal Geffen; Data Acquisition; Florian Schlagenhauf: Data acquisition. Fares Alouf: Data acquisition. Stephan Eliez: Review and editing of manuscript. Stefan Kaiser: Resources, Review and editing of manuscript. Indrit Bègue: Conceptualization, Investigation, Resources, Supervision, Project administration, Funding acquisition, Writing original manuscript.

Farnaz Delavari and Jade Awada contributed equally to this article.

## Conflicts of interest

SK has received advisory board honoraria from Boehringer Ingelheim and Exeltis. The other authors report no conflicts of interests.

## Notes

### Author Declarations

Geneva Ethics Committee in Switzerland gave approval for this work (CCER. BASEC ID: 2017-01765 and 2020-02169).

### Summary of Updates

We conducted additional analyses using an independently acquired sample of individuals with schizophrenia. Remarkably, we observed the same results as in our primary cohort, strengthening the robustness of our findings. Here are the 3 key highlights of our work: 1)Identification of a reproducible cerebellum VTA anticoactivation pattern (r = 0.98) in bilateral paravermal Crus I/II across two distinct recordings at baseline and 3 months (Cohort in Geneva, Switzerland). 2)Clinical relevance of this pattern: lower anticoactivation emergence (i.e., how often the pattern is encountered) at baseline correlated with more severe apathy, particularly asociality and avolition. At 3 months, lower anticoactivation persistence (i.e., how long the pattern lasted) was associated with greater apathy, especially anhedonia, and predicted increased anhedonia at 9 months. Similarly, reduced anticoactivation emergence at 3 months predicted worse asociality at 9 months while reduced persistence at 3 months related to worse anhedonia at 9 months 3)Validation in an independent cohort (Cohort in Berlin, Germany): we identify this pattern with a spatial similarity to the pattern identified in the main cohort of r = 0.93 and we replicate the association of the pattern emergence with apathy, particularly asociality. Please refer to the Validation dataset analyses part. Figures are updated: Former Figure 1 has been moved to the Supplemental Materials. Figure 2 is now Figure 1, with updates to panels B and C. Figure 3 is now Figure 2, with updates to panels C and D. Figure 4 is now Figure 3, with an added panel B. A new Figure 4 has been created to illustrate the additional analyses on validation cohort. Supplemental materials are also updated, with additional analyses.

