## Supplemental Materials for "A Longitudinal and Reproducible Anti-coactivation Pattern Between the Cerebellum and the Ventral Tegmental Area Relates to Apathy in Schizophrenia"

1. **Clinical psychopathological and cognitive assessment**

All individuals (HC and SZ) underwent a detailed psychopathological and cognitive assessment. To assess the negative symptoms in patients with SZ, we used the Brief Negative Symptoms Scale (BNSS)(1), a semi-structured interview conducted by trained researchers/clinicians. The BNSS quantitatively measures negative symptoms across five individual symptoms (anhedonia, asociality, avolition, flat affect, alogia). BNSS items are scored on a 7-point severity scale, higher scores indicate more severe impairment. The BNSS is recognized for its validity, reliability, and sensitivity, rendering it an optimal instrument for our investigation. Additionally, we employed the Positive and Negative Syndrome Scale (PANSS)(2), the Calgary Depression Scale for Schizophrenia (CDS)(3), and the Personal and Social Performance Scale (PSP)(4). Cognitive function was assessed with the Brief Assessment of Cognition in Schizophrenia (BACS) battery. The BACS Battery is intended to provide a relatively brief evaluation of cognitive function in schizophrenia (5). Extrapyramidal syndrome was assessed with St. Hans scale (6). Antipsychotic medication (in risperidone equivalents) was also recorded.

2. **Exclusion criteria**

Patients were not included if they presented other Axis I disorders, in particular, comorbid major depressive episodes, florid psychotic symptoms (Positive and Negative Syndrome Scale, any positive item > 4 (2)), significant extrapyramidal side effects (any St Hans Rating scale item > 3 (6)), use of benzodiazepines or lorazepam equivalents of more than 1mg, or active use of substances (including opiate substitution for medical purposes). Healthy controls were included only if they did not present any Axis I disorder, a family history of psychotic disorders or psychotropic drug use.

3. **Neuroimaging sequence parameters**

**Main cohort**

MRI data were acquired at the Campus Biotech (Geneva, Switzerland) using a Siemens 3T Magnetom Prisma scanner and a standard 64-channel head coil. High-resolution structural images were captured utilizing a T1-weighted sequence with specified parameters (voxel size = 1.0 × 1.0 × 1.0 mm^3^, repetition time = 2200 ms, echo time = 2.96 ms, field of view read = 256 mm, flip angle = 9°, slice thickness = 1.0 mm, phase encoding: anterior > posterior, no fat suppression) to ensure optimal volumetric resolution. Resting-state functional MRI (rs-fMRI) scans were acquired with the following parameters: 589 frames (9.8 minutes), field of view read = 224 mm, voxel size = 2.0 × 2.0 × 2.0 mm^3^, 66 transversal slices, slice thickness = 2.0 mm, repetition time = 1000 ms, echo time = 32.0 ms, flip angle = 50°, phase encoding: anterior > posterior, acceleration mode with factor for parallel imaging = 6.

**Validation cohort**

Validation cohort data were acquired with a Siemens Magnetom Prisma Fit 3T scanner at Charité Hospital in Berlin with a 64-channel head coil with the same acquisition protocol as the main cohort.

4. **Preprocessing**

The pipeline included initial quality assessment followed by realignment of functional images, spatial normalization, and smoothing using a 6 mm full width at half maximum Gaussian kernel. Additionally, structural images were co-registered and segmented, using Diffeomorphic Anatomical Registration Through Exponentiated Lie algebra (DARTEL). The 5 initial functional scans were excluded to allow for signal stabilization. Linear detrending was performed and nuisance covariates (constant, linear and quadratic trends, the six head movement parameters, mean white matter and cerebrospinal fluid time courses) were regressed from the BOLD signal. White matter and cerebrospinal fluid signals were located using conservative tissue masks from the DPARSF toolbox. Ultimately, images were normalized to the DARTEL and subsequently MNI spaces for group-level analysis. Given the significant group difference in frame-wise displacement, extended motion correction was done in the form of scrubbing high-motion frames later within the analysis (see Table 1 for a summary of mean framewise displacement across groups).

5. **Quality check**

**Main cohort**

In the main cohort, 32 patients and 9 controls at T1 and 14 patients and 10 controls at T2 were excluded during different stages of quality control of the imaging analysis. Out of 33 patients who returned for interviews at T3, only 21 had scans in T2. Thus, the final main cohort consisted of 105 individuals (58 SZ and 47 HC) at T1, 41 individuals (22 SZ and 19 HC) at T2, and 21 SZ patients at T3.

At T1 and among the excluded subjects, 11 lacked the resting-state scan, 3 patients and 4 healthy controls moved excessively above the threshold of 3 mm, and 8 participants missed parts of the cerebellum within the field of view. During extended motion correction, 7 patients and 4 healthy controls lost more than half of their scanned frames due to scrubbing of the frames with framewise displacement higher than 0.5 mm, and therefore were excluded from CAP analysis. At T2, 14 patients and 10 healthy controls were excluded from imaging analysis. Among those, 15 lacked the resting-state scan, 3 moved excessively above the threshold of 3 mm, 5 interrupted the study, 1 was excluded because of epileptic seizure and 1 missed parts of the cerebellum within the field of view. At T3, among the 33 participants who returned for interviews, 12 patients were excluded from the analysis because they were already excluded from the imaging analysis at T2 (see Table S1).

**Table S1: Screening process and participants’ data retention in the main cohort at baseline (T1), 3-month follow-up (T2) and 9-month follow-up (T3), as well as in the validation cohort. Abbreviations: SZ = schizophrenia; HC = healthy controls; NA: not available.**

| **Main cohort** | | **Validation cohort** | |
| --- | --- | --- | --- |
| *Screening* | | | |
| 333 SZ | 94 HC | 141 SZ | 62 HC |
| *Number of participants recruited* | | | |
| 87 SZ | 55 HC | 47 SZ | 36 HC |
| *Number of participants lacking resting-state scans* | | | |
| 11 | | 8 SZ | 1 HC |
| *Number of participants excluded after quality check* | | | |
| 18 SZ | 8 HC | 11 SZ | 3 HZ |
| *Number of participants included at T1* | | | |
| 58 SZ | 47 HC | 28 SZ | 25 HC |
| *Number of participants who returned at T2* | | | |
| 36 SZ | 29 HC | NA | |
| *Number of participants excluded after quality check* | | | |
| 14 SZ | 10 HC | NA | |
| *Number of participants included at T2* | | | |
| 22 SZ | 19 HC | NA | |
| *Number of participants who returned at T3* | | | |
| 33 SZ | NA | NA | |
| *Number of participants excluded after quality check and/or previously excluded at T2* | | | |
| 12 SZ | NA | NA | |
| *Number of participants included at T3* | | | |
| 21 SZ | NA | NA | |
| *Percentages of drop out from T1 and T2:* *30% (HC), 40% (SZ).* | | | |

To ensure the absence of selection bias, we compared psychosis ratings, medication levels and negative symptoms scores between patients excluded due to movement and those included in CAP analysis. No significant differences were found in any score. Results are shown in Table S2.

| **Table S2. Comparison of psychosis ratings, medication and negative symptoms scores between patients with schizophrenia included in CAP analysis (SZ included) and those excluded due to movement (SZ excluded) in the main cohort. Standard deviations are provided in parentheses. Abbreviations: SZ = schizophrenia; df = degrees of freedom; PANSS = Positive and Negative Symptom Scale; BNSS = Brief Negative Symptoms Score.** | | | | | | | |
| --- | --- | --- | --- | --- | --- | --- | --- |
|  | **SZ included** | **SZ excluded** |  | **Sum of squares** | **df** | **F** | ***p*** |
| **PANSS psychosis** | 9.488 (2.634) | 10.125 (3.243) | **Group** | 5.419 | 1 | 0.722 | 0.398 |
|  |  |  | **Residual** | 705.738 | 94 |  |  |
| **BNSS total** | 19.288 (14.569) | 21.688 (12.387) | **Group** | 76.8 | 1 | 0.379 | 0.54 |
|  |  |  | **Residual** | 19069.825 | 94 |  |  |
| **BNSS anhedonia** | 4.263 (4.867) | 6 (3.578) | **Group** | 40.252 | 1 | 1.833 | 0.179 |
|  |  |  | **Residual** | 2063.488 | 94 |  |  |
| **BNSS asociality** | 3.575 (2.589) | 3.813 (2.588) | **Group** | 0.752 | 1 | 0.112 | 0.738 |
|  |  |  | **Residual** | 629.988 | 94 |  |  |
| **BNSS avolition** | 4.338 (2.959) | 4 (2.658) | **Group** | 1.519 | 1 | 0.179 | 0.673 |
|  |  |  | **Residual** | 797.888 | 94 |  |  |
| **BNSS flat affect** | 4.825 (4.331) | 6.063 (3.907) | **Group** | 20.419 | 1 | 1.1222 | 0.292 |
|  |  |  | **Residual** | 1710.488 | 94 |  |  |
| **BNSS apathy** | 12.175 (8.783) | 13.813 (7.213) | **Group** | 35.752 | 1 | 0.489 | 0.486 |
|  |  |  | **Residual** | 6873.988 | 94 |  |  |
| **BNSS alogia** | 1.925 (3.240) | 1.375 (2.306) | **Group** | 4.033 | 1 | 0.417 | 0.52 |
|  |  |  | **Residual** | 909.3 | 94 |  |  |
| **BNSS diminished expression** | 6.75 (7.041) | 7.438 (5.621) | **Group** | 6.302 | 1 | 0.135 | 0.714 |
|  |  |  | **Residual** | 4390.938 | 94 |  |  |
| **Medication (dose in risperidone equivalent)** | 4.485 (2.555) | 5.264 (2.492) | **Group** | 8.066 | 1 | 1.245 | 0.267 |
|  |  |  | **Residual** | 608.835 | 94 |  |  |

**Validation cohort**

In the validation cohort, 83 individuals (47 SZ and 36 HC) were recruited. 19 patients and 11 controls were excluded during different stages of quality control of the imaging analysis. The final validation cohort comprised 53 individuals (28 SZ and 25 HC).

A summary of the reasons for exclusion and percentages of drop-out are detailed in Table S1.

Among the excluded subjects, 11 patients lost more than half of their scanned frames due to scrubbing of the frames with framewise displacement higher than 0.5 mm. Resting-state scans were missing for 10 patients and 1 control, while 3 controls had incomplete scans.

To ensure the absence of selection bias, we also compared psychosis ratings, medication levels and negative symptoms scores between patients excluded due to movement and those included in CAP analysis. No significant differences were found in any score. Results are shown in Table S3.

| **Table S3. Comparison of psychosis ratings, medication and negative symptoms scores between patients with schizophrenia included in CAP analysis (SZ included) and those excluded due to movement (SZ excluded) in the validation cohort. Standard deviations are provided in parentheses. Abbreviations: SZ = schizophrenia; df = degrees of freedom; PANSS = Positive and Negative Symptom Scale; BNSS = Brief Negative Symptoms Score.** | | | | | | | |
| --- | --- | --- | --- | --- | --- | --- | --- |
|  | **SZ included** | **SZ excluded** |  | **Sum of squares** | **df** | **F** | ***p*** |
| **PANSS psychosis** | 12.964 (5,548) | 9.9 (4.795) | **Group** | 69.188 | 1 | 2.4 | 0.131 |
|  |  |  | **Residual** | 1037.864 | 36 |  |  |
| **BNSS total** | 23.929 (15.993) | 27.2 (16.349) | **Group** | 78.859 | 1 | 0.305 | 0.584 |
|  |  |  | **Residual** | 9311.457 | 36 |  |  |
| **BNSS anhedonia** | 6.536 (4.574) | 8 (4.472) | **Group** | 15.79887 | 1 | 0.763 | 0.388 |
|  |  |  | **Residual** | 744.9643 | 36 |  |  |
| **BNSS asociality** | 4.25 (2.648) | 4 (3.018) | **Group** | 0.461 | 1 | 0.061 | 0.806 |
|  |  |  | **Residual** | 271.25 | 36 |  |  |
| **BNSS avolition** | 4.25 (2.837) | 5.6 (2.066) | **Group** | 13.429 | 1 | 1.891 | 0.178 |
|  |  |  | **Residual** | 255.65 | 36 |  |  |
| **BNSS flat affect** | 5.179 (4.643) | 6.1 (4.795) | **Group** | 6.256 | 1 | 0.285 | 0.596 |
|  |  |  | **Residual** | 789.0071 | 36 |  |  |
| **BNSS apathy** | 15.036 (9.343) | 16 (8.628) | **Group** | 6.852 | 1 | 0.081 | 0.777 |
|  |  |  | **Residual** | 3026.964 | 36 |  |  |
| **BNSS alogia** | 1.929 (2.981) | 2.2 (2.394) | **Group** | 0.543 | 1 | 0.067 | 0.797 |
|  |  |  | **Residual** | 291.458 | 36 |  |  |
| **BNSS diminished expression** | 7.107 (6.973) | 10.6 (6.67) | **Group** | 89.895 | 1 | 1.890 | 0.178 |
|  |  |  | **Residual** | 1713.079 | 36 |  |  |
| **Medication (dose in risperidone equivalent)** | 6.152 (4.331) | 5.110 (3.613) | **Group** | 8.004 | 1 | 0.462 | 0.501 |
|  |  |  | **Residual** | 623.986 | 36 |  |  |

6. **Co-activation pattern (CAP) analysis**

Neural activity during rest is organized into time-varying, recurrent patterns of cross-regional interactions that can be captured via dynamic resting-state functional connectivity approaches (7–9). To examine dynamic functional connectivity between the cerebellum and the VTA, we leveraged coactivation pattern (CAP) analysis (10,11), an established approach that has already contributed to the study of various psychiatric disorders (12,13) and implemented in the publicly available *TbCAPs* toolbox (14).

This method has been applied across different research fields:

- *Liu and Duyn, 2013 (10):* they proposed clustering selected individual BOLD volumes of a resting-state scan based on spatial similarity (“coactivation patterns”, or CAPs).
- *Liu et al., 2018 (11)*: they discussed the relevance of CAPs and reviewed the development and recent advances of this methodology.
- *Kaiser et al*., 2019 (12): they studied fronto-insular-default network dynamics in adolescents with depression and rumination.
- *Huang et al., 2020 (15)*: they used CAPs to investigate human consciousness and its dynamic brain activity.
- *Goodman et al*., 2021 (16): they examined relationships between functional brain dynamics and subclinical-to-mild depressive symptomatology in a large community sample of adults with and without psychiatric diagnoses.
- *Yang et al*., 2021 (17): they investigated the reproducibility and generalizability of CAP analysis across different methodological pipelines and cohorts and identified reliable CAP states and their dynamic characteristics, providing information about aberrant brain dynamic configurations for understanding the psychopathological mechanisms in schizophrenia.
- *Delavari et al., 2024 (18)*: they studied the temporal variability of the thalamus in functional networks in adult patients with schizophrenia (22q11.2 deletion syndrome).
- *An et al., 2024 (19)*: they investigated spatial and temporal dynamics in patients with major depressive disorder (MDD) using coactivation pattern analysis.

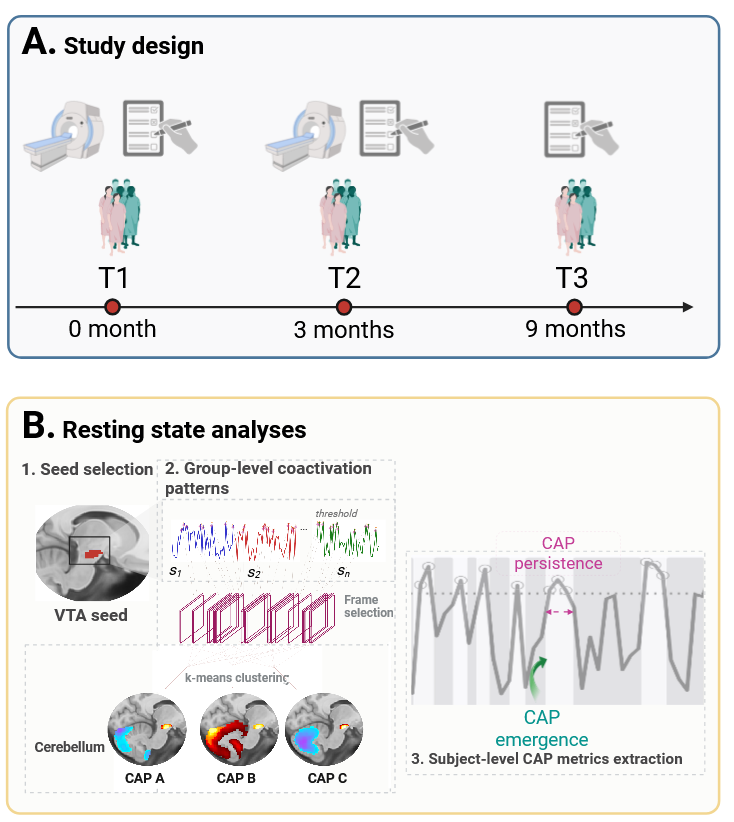

**Figure S1.** **Study design and coactivation pattern (CAP) analysis overview.** A. Study design. Main cohort: healthy controls (HC) and patients with schizophrenia (SZ) underwent an MRI scan and detailed psychopathological assessment at baseline (T1) and at 3 months of follow-up (T2). At 9 months after baseline (T3), only patients were invited and assessed with psychopathological testing alone. Validation cohort: identical evaluation as T1 baseline was performed for HC and SZ patients (not shown in figure). B. Resting-state analysis. B.1-2. Average VTA signal (seed) for each individual is standardized, thresholded, and timepoints exceeding this threshold (red circles) are selected, denoting timepoints of prominent VTA activity. We then performed a *K*-means clustering on the cerebellar voxels at these timepoints to obtain cerebellar patterns reflective of coactivation with the VTA. B.3. Within each individual, for each CAP, persistence is calculated as the average time in seconds a CAP lasts after occurring. CAP emergence is measured as the number of times a specific CAP occurred after episodes of VTA under-threshold activity.

7. **Selection of the number of clusters for the CAP analysis**

**Main cohort**

The number of clusters for *K*-means clustering was chosen following a train-and-test procedure: first, subjects were randomly split into half to form test and training sets. Both sets of subjects went through frame selection based on activation of the VTA, resulting in the test and training sets of frames. We then ran *K*-means clustering on the training frames for different candidate numbers of clusters (*K* = 2 to 10). Each *K*-means clustering resulted in *K* training centroids. Subsequently, the test frames were assigned to the training centroids for which they showed the lowest distance, as calculated by 1-correlation – the same distance measure as used in the *K*-means clustering algorithm. The sum of the distances (sumD) of test frames to the training centroids was calculated for each of the *K* centroids. For each candidate *K* value, the cluster with the highest sum of distances was then the cluster with the worst fit. The sum of distances for this worst-fit cluster (maximum sumD) was monitored across candidate *K* values (2 to 10). The *K* with lower sumD would indicate a good solution for the number of clusters as the distance of data points to the centroid is minimal even for the worst cluster. We repeated this procedure for 20 folds of random splitting across the subjects. Figure S1 displays the maximum sumD for all candidate K values across all 20 folds. *K* = 4 shows low maximum sumD with concomitantly narrow standard deviation across folds, hence representing a good solution with high reproducibility across subjects.

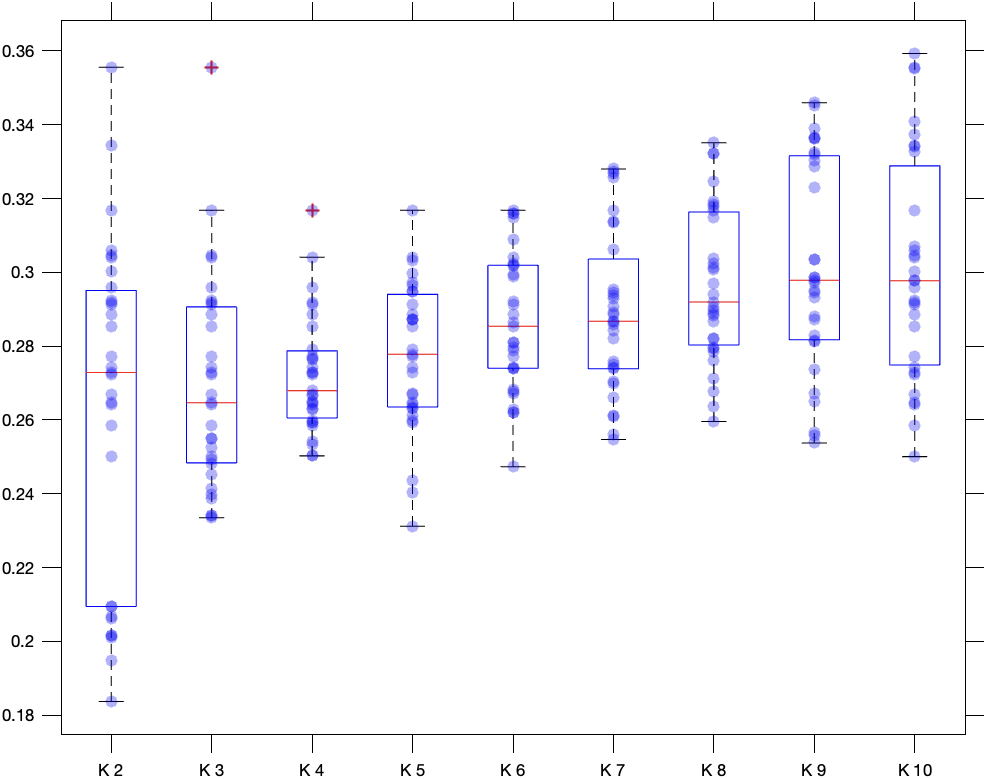

**Figure S2. Sum of distances for the cluster with maximum sum of distances in the train-and-test procedure (main cohort).**

**Validation cohort**

We followed the same procedure with the validation cohort and selected *K* = 7 clusters as an optimum, since maximum sumD remained lower than expected with respect to the trend seen with increasing *K*, denoting particularly satisfying clustering at this granularity.

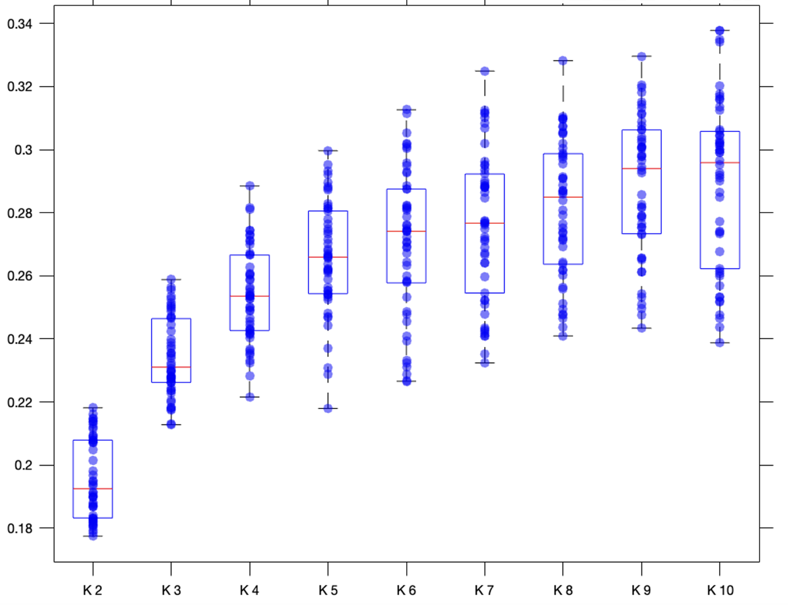

**Figure S3. Sum of distances for the cluster with maximum sum of distances in the train-and-test procedure (validation cohort).**

8. **Matching matrix between CAPs**

We computed Pearson correlations between all CAP pairs at T1 and T2, organizing the results into a correlation matrix. Scatter plots of T1 vs. T2 CAP1 voxel-wise signal values for the HC and SZ groups were generated separately (Figure S3). Additionally, we plotted CAP1 signal values at T1 in the main cohort against those in the validation cohort (Figure S4).

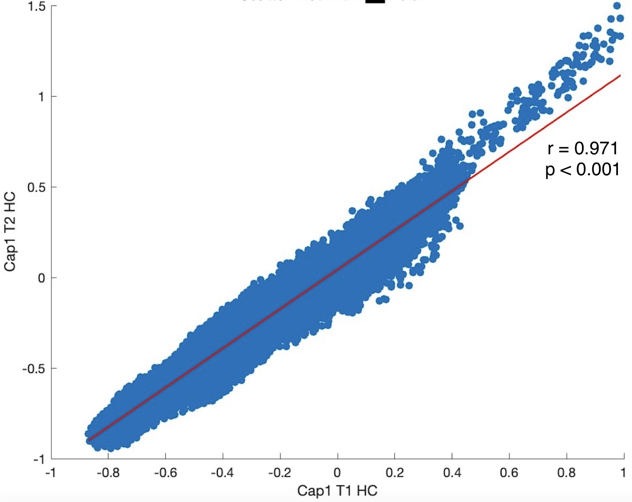

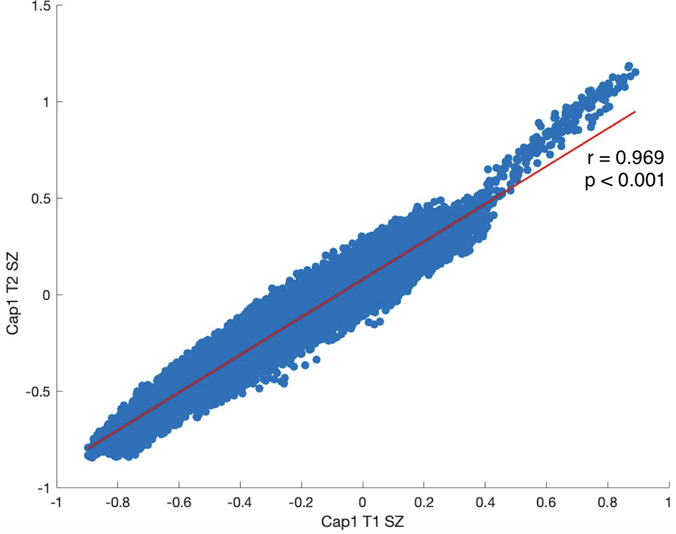

**Figure S4. Scatter plots of CAP1 signal values contrasted between T1 and T2 in HC (on the left) and in SZ patients (on the right) in the main cohort.**

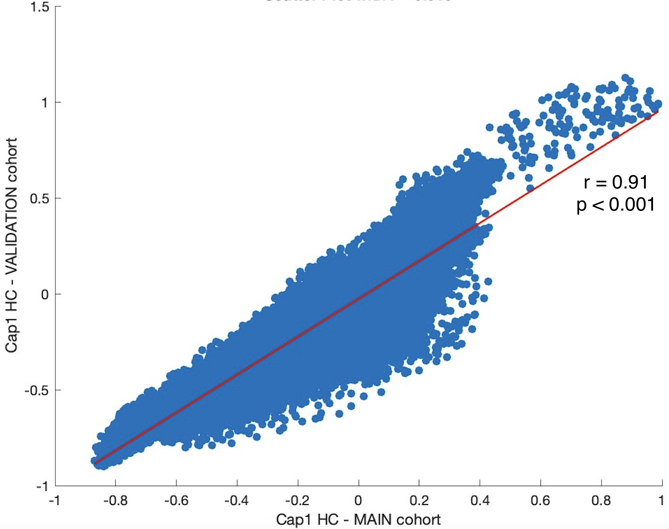

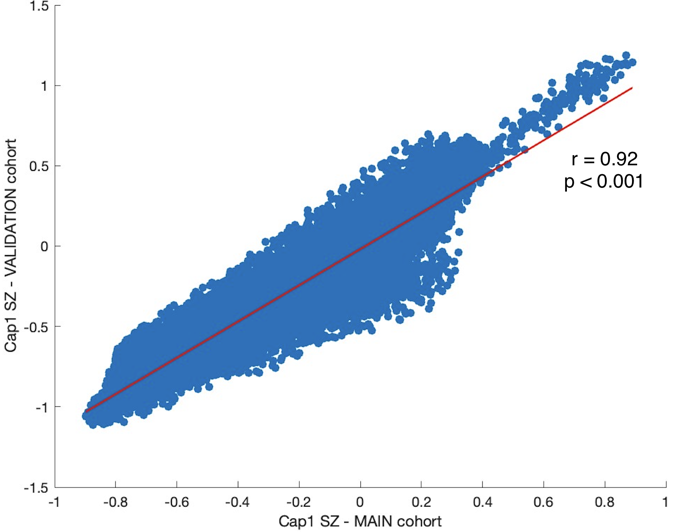

**Figure S5. Scatter plot of CAP1 signal values contrasted between T1 in the main cohort and the validation cohort, shown for HC (on the left) and in SZ patients (on the right).**

9. **Longitudinal psychopathology analyses**

One-way ANOVAs were performed to longitudinally assess the negative symptoms of schizophrenia (BNSS), as well as psychosis (PANSS), depression (CDS), psychosocial functioning (PSP), anti-psychotic medication (dose in risperidone equivalent), extrapyramidal syndrome (SHRS) and cognition (BACS). These results are reported in Table S4.

The validation cohort scores were also included in the ANOVAs. We found a significant difference in PANSS psychosis scores between the two cohorts (see Table S5 for post-hoc analysis, uncorrected for multiple comparisons).

**Table S4. Summary of means (with standard deviations) for selected variables of the main cohort at T1, T2, T3, as well as the validation cohort. Abbreviations: SD = standard deviation; df = degrees of freedom; BNSS = Brief Negative Symptoms Score; dimex = diminished expression; PANSS = Positive and Negative Symptom Scale; CDS = Calgary Depression Scale for Schizophrenia; PSP = Personal and Social Performance Scale; risp = dose in risperidone equivalent; SHRS = St. Hans Rating Scale; BACS = Brief Assessment of Cognition in Schizophrenia**

|  | **T1** | | **T2** | | **T3** | | **Validation cohort** | |  | **Sum of Squares** | **df** | **F** | ***p*** |
| --- | --- | --- | --- | --- | --- | --- | --- | --- | --- | --- | --- | --- | --- |
|  | **Mean** | **SD** | **Mean** | **SD** | **Mean** | **SD** | **Mean** | **SD** |  |  |  |  |  |
| **BNSS total** | 19.603 | 14.573 | 18.454 | 14.867 | 17.476 | 13.761 | 23.929 | 15.993 | **Time** | 627.5709 | 3 | 0.953 | 0.417 |
|  |  |  |  |  |  |  |  |  | **Residual** | 27440.43 | 125 |  |  |
| **BNSS anhedonia** | 4.327 | 4.766 | 4.09 | 5.236 | 4.143 | 5.275 | 6.536 | 4.574 | **Time** | 116.909 | 3 | 1.628 | 0.186 |
|  |  |  |  |  |  |  |  |  | **Residual** | 2992.13 | 125 |  |  |
| **BNSS asociality** | 3.862 | 2.558 | 2.818 | 2.575 | 2.762 | 2.211 | 4.25 | 2.648 | **Time** | 43.996 | 3 | 2.294 | 0.081 |
|  |  |  |  |  |  |  |  |  | **Residual** | 799.2288 | 125 |  |  |
| **BNSS avolition** | 4.276 | 3.065 | 4.5 | 2.721 | 4.190 | 3.234 | 4.25 | 2.837 | **Time** | 1.232 | 3 | 0.046 | 0.987 |
|  |  |  |  |  |  |  |  |  | **Residual** | 1117.574 | 125 |  |  |
| **BNSS flat affect** | 4.741 | 4.459 | 5.045 | 4.06 | 4.857 | 3.511 | 5.179 | 4.643 | **Time** | 4.130 | 3 | 0.075 | 0.974 |
|  |  |  |  |  |  |  |  |  | **Residual** | 2308.754 | 125 |  |  |
| **BNSS alogia** | 1.914 | 3.180 | 1.954 | 3.471 | 1.381 | 2.801 | 1.929 | 2.981 | **Time** | 5.248 | 3 | 0.178 | 0.911 |
|  |  |  |  |  |  |  |  |  | **Residual** | 1226.333 | 125 |  |  |
| **BNSS dimex** | 6.655 | 7.097 | 7 | 7.051 | 6.238 | 5.761 | 7.107 | 6.973 | **Time** | 10.967 | 3 | 0.078 | 0.972 |
|  |  |  |  |  |  |  |  |  | **Residual** | 5891.592 | 125 |  |  |
| **BNSS apathy** | 12.466 | 8.672 | 11.409 | 9.231 | 11.095 | 8.927 | 15.036 | 9.343 | **Time** | 245.864 | 3 | 1.022 | 0.385 |
|  |  |  |  |  |  |  |  |  | **Residual** | 10026.52 | 125 |  |  |
| **PANSS psychosis** | 9.603 | 2.655 | 9.182 | 2.612 | 9.762 | 3.192 | 12.964 | 5.548 | **Time** | 260.462 | 3 | 6.869 | **0.0002** |
|  |  |  |  |  |  |  |  |  | **Residual** | 1579.926 | 125 |  |  |
| **CDS** | 2.19 | 3.081 | 2.318 | 3.168 | 1.667 | 2.415 | 2.107 | 2.78 | **Time** | 5.449 | 3 | 0.211 | 0.889 |
|  |  |  |  |  |  |  |  |  | **Residual** | 1077.032 | 125 |  |  |
| **PSP** | 4.397 | 1.736 | 5.091 | 1.444 | 5.095 | 1.758 | 4.964 | 1.895 | **Time** | 13.591 | 3 | 1.512 | 0.215 |
|  |  |  |  |  |  |  |  |  | **Residual** | 374.471 | 125 |  |  |
| **risp** | 4.509 | 2.629 | 4.42 | 2.407 | 4.652 | 2.814 | 6.152 | 4.331 | **Time** | 58.993 | 3 | 2.082 | 0.106 |
|  |  |  |  |  |  |  |  |  | **Residual** | 1180.434 | 125 |  |  |
| **SHRS** | 1.017 | 0.827 | 1.136 | 1.246 | 1.143 | 1.195 | 1.5 | 3.727 | **Time** | 4.436 | 3 | 0.389 | 0.761 |
|  |  |  |  |  |  |  |  |  | **Residual** | 475.145 | 125 |  |  |
| **BACS** | 35.672 | 10.743 | 33.955 | 13.185 | 37.762 | 11.287 | 38.25 | 13.01 | **Time** | 295.164 | 3 | 0.706 | 0.55 |
|  |  |  |  |  |  |  |  |  | **Residual** | 17410.79 | 125 |  |  |

**Table S5. Post-hoc analysis (Tukey’s honestly significant difference test, uncorrected for multiple comparisons) on PANSS psychosis scores in the main cohort at baseline (T1), 3 months of follow-up (T2), and 9 months of follow-up (T3) and in the validation cohort (B1).**

| **Group 1** | **Group 2** | **Mean difference (Group 2 – Group 1)** | ***p*** | **95% confidence interval** | |
| --- | --- | --- | --- | --- | --- |
|  |  |  |  | **Lower bound** | **Upper bound** |
| B1 | T1 | -3.361 | 0.0004 | -5.4912 | -1.2305 |
| B1 | T2 | -3.783 | 0.0016 | -6.4199 | -1.145 |
| B1 | T3 | -3.202 | 0.0119 | -5.8748 | -0.53 |
| T1 | T2 | -0.422 | 0.965 | -2.7396 | 1.8964 |
| T1 | T3 | 0.159 | 0.998 | -2.1992 | 2.5161 |
| T2 | T3 | 0.580 | 0.950 | -2.2442 | 3.4044 |

10. **Correlation between negative symptoms and other clinical variables**

To examine whether negative symptoms related to other clinical variables, we computed Spearman correlations between negative symptoms (apathy and diminished expression) and selected variables such as medication (dose in risperidone equivalent), cognition (bac_total_t), psychosis (panspos), depression (cdc_total_Depression), psychosocial functioning (psp_total_psychologicalfunctioning), and extrapyramidal syndrome (shr_parkinson_global) at T1, T2 and T3.

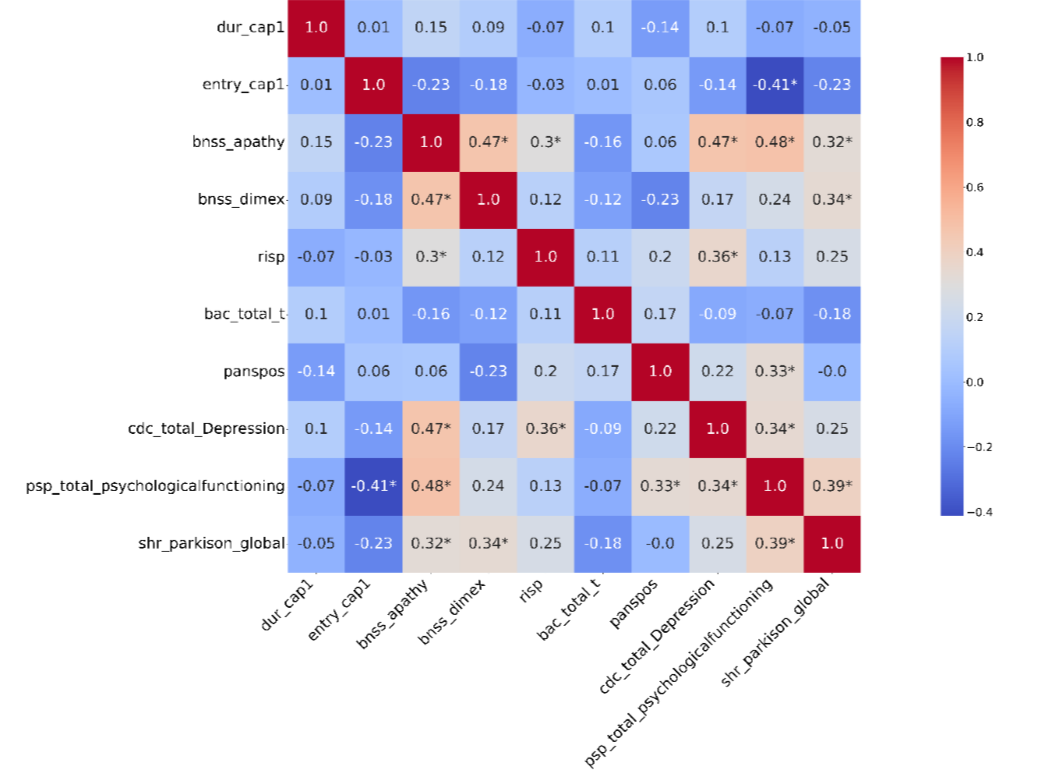

**Figure S6. Spearman correlation matrix between CAP1 persistence (*i.e.*, duration) and emergence (*i.e.*, entry) with selected clinical variables at T1.**

*Note: selected variables are BNSS scores (apathy and diminished expression), medication (risp), cognition (bac_total_t), psychosis (panspos), depression (cdc_total_Depression), psychosocial functioning (psp_total_psychologicalfunctioning), and extrapyramidal syndrome (shr_parkinson_global).*

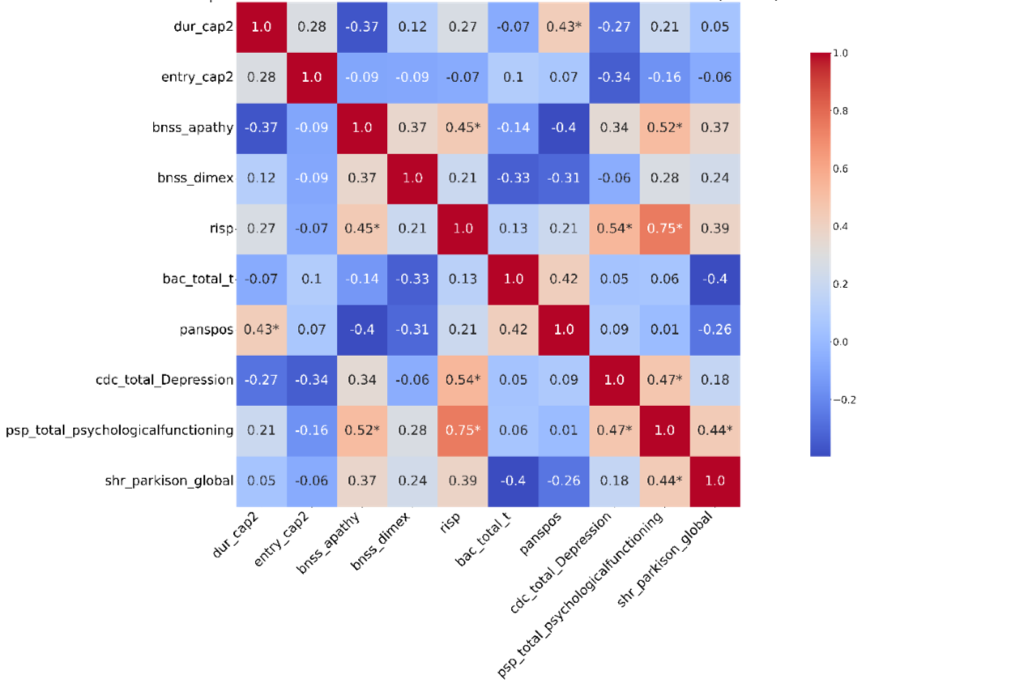

**Figure S7. Spearman correlation matrix between CAP1 persistence (*i.e.*, duration) and emergence (*i.e.*, entry) with selected clinical variables at T2.**

*Note: selected variables are BNSS scores (apathy and diminished expression), medication (risp), cognition (bac_total_t), psychosis (panspos), depression (cdc_total_Depression), psychosocial functioning (psp_total_psychologicalfunctioning), and extrapyramidal syndrome (shr_parkinson_global).*

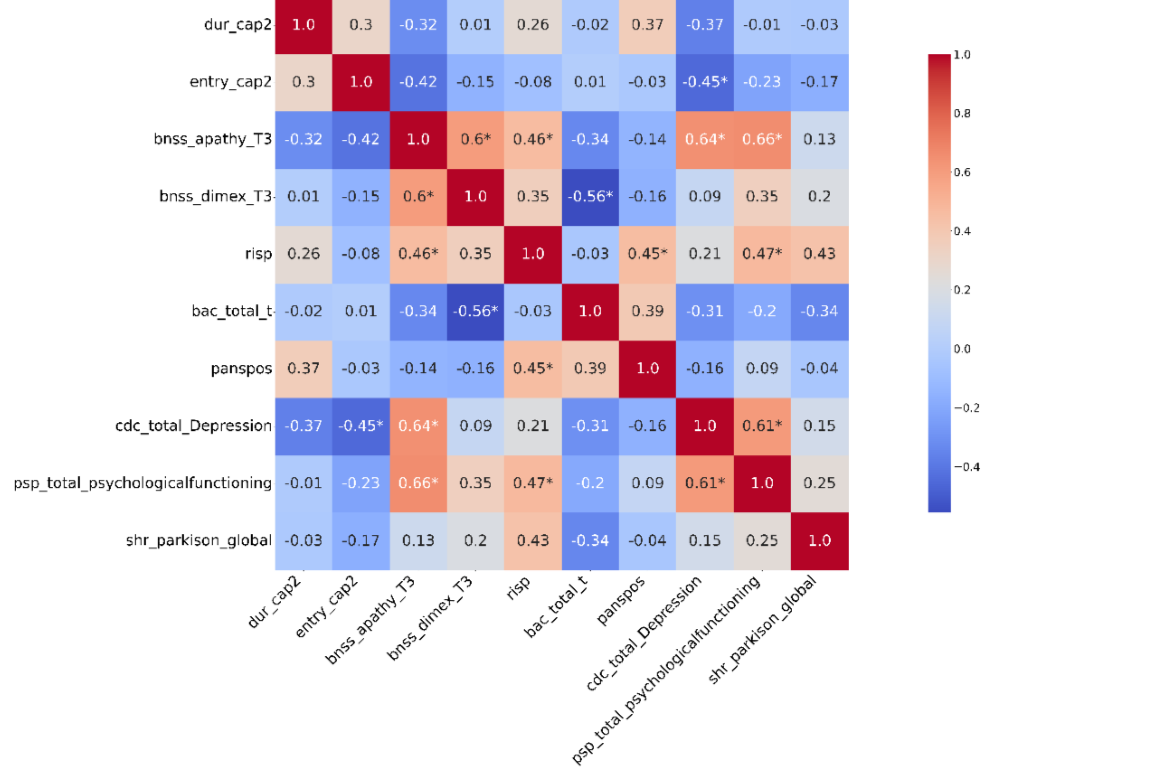

**Figure S8. Spearman correlation matrix between CAP1 persistence (*i.e.*, duration) and emergence (*i.e.*, entry) with selected clinical variables at T3.**

*Note: selected variables are BNSS scores (apathy and diminished expression), medication (risp), cognition (bac_total_t), psychosis (panspos), depression (cdc_total_Depression), psychosocial functioning (psp_total_psychologicalfunctioning), and extrapyramidal syndrome (shr_parkinson_global).*

11. **Non-parametric permutation tests on partial correlations**

To evaluate the statistical significance of partial correlations, a non-parametric permutation testing approach was also employed. The observed partial correlation between variables was first calculated, controlling for covariates, using Spearman's rank correlation. Under the null hypothesis of no association between the variables, the data were permuted 5’000 times. Specifically, one variable in the pair was randomly shuffled while keeping the other variable and covariates fixed, thereby breaking the original relationship. For each permutation, the partial correlation was recalculated to generate a null distribution of correlations expected under the null hypothesis. The p-value was then calculated as the proportion of permuted correlations that were as extreme as or more extreme than the observed correlation, using a two-tailed test. This method makes no assumptions about the underlying data distribution, ensuring robustness for non-normally distributed or small sample datasets. A threshold of p<0.05 was used to determine statistical significance.

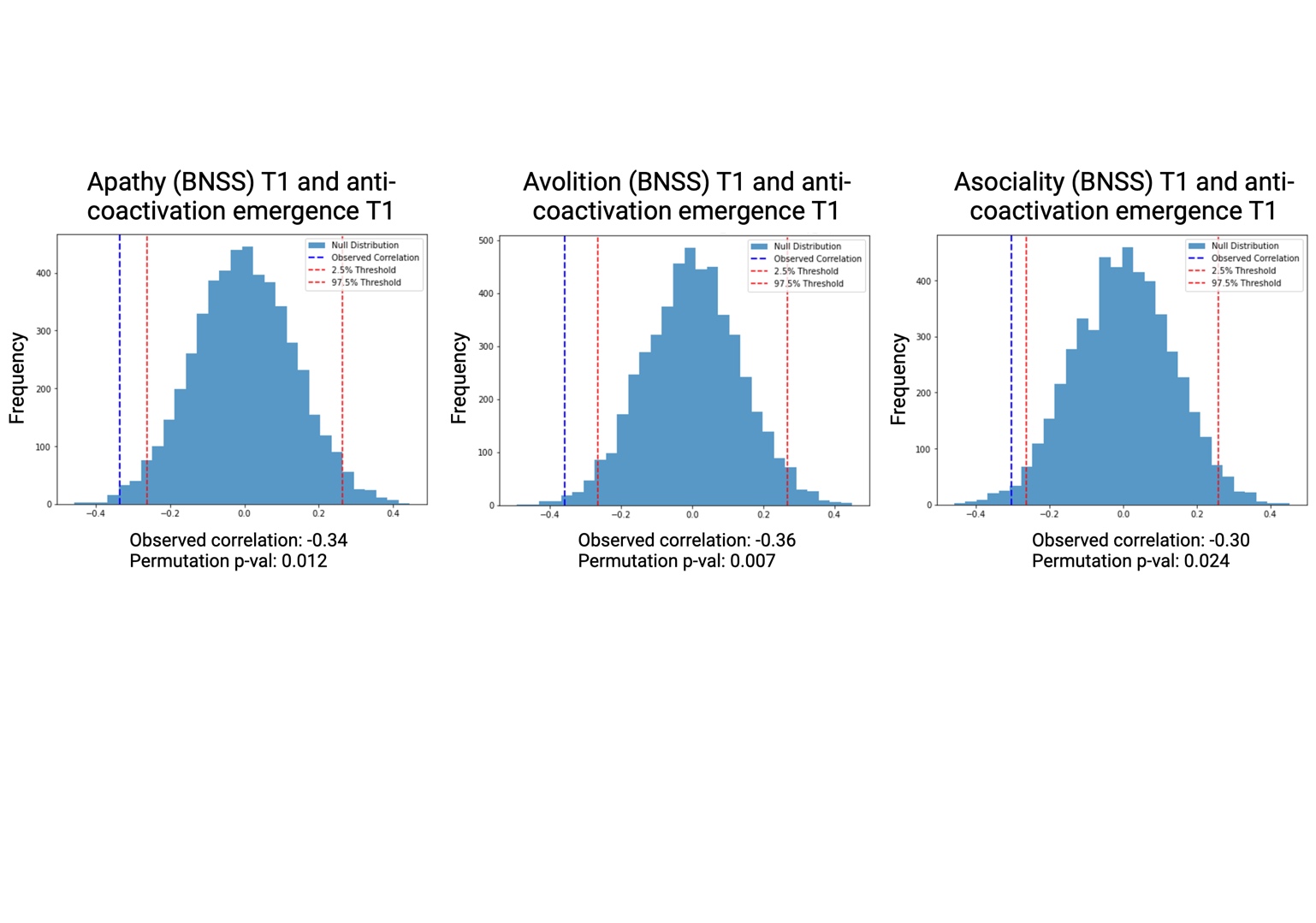

**Figure S9. Main cohort: non-parametric permutation tests at T1** **between CAP1 emergence (CAP1 entry frequency) and BNSS apathy, avolition, and asociality.**

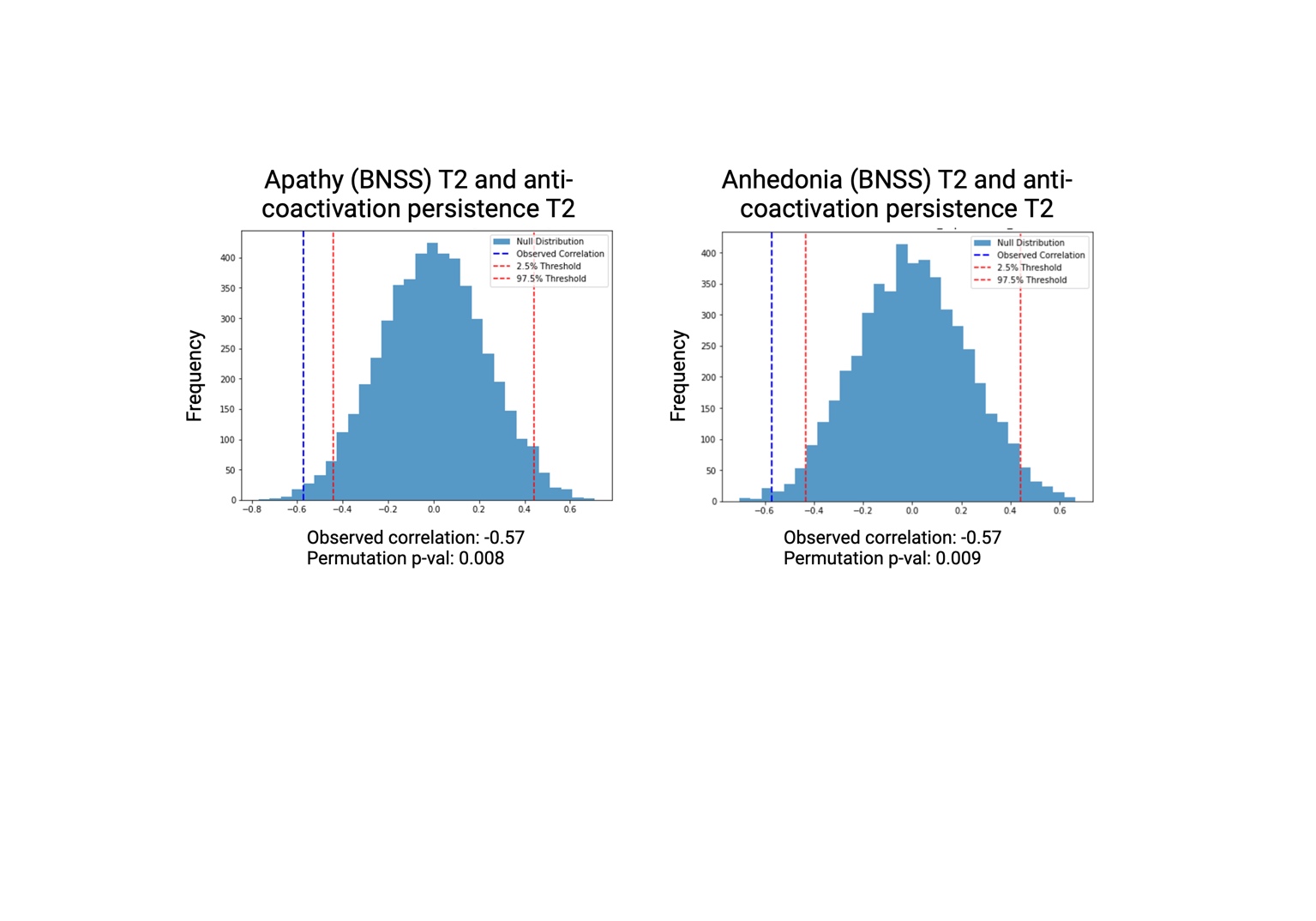

**Figure S10. Main cohort: non-parametric permutation tests at T2** **between CAP1 persistence (CAP1 duration in seconds) and BNSS apathy and anhedonia.**

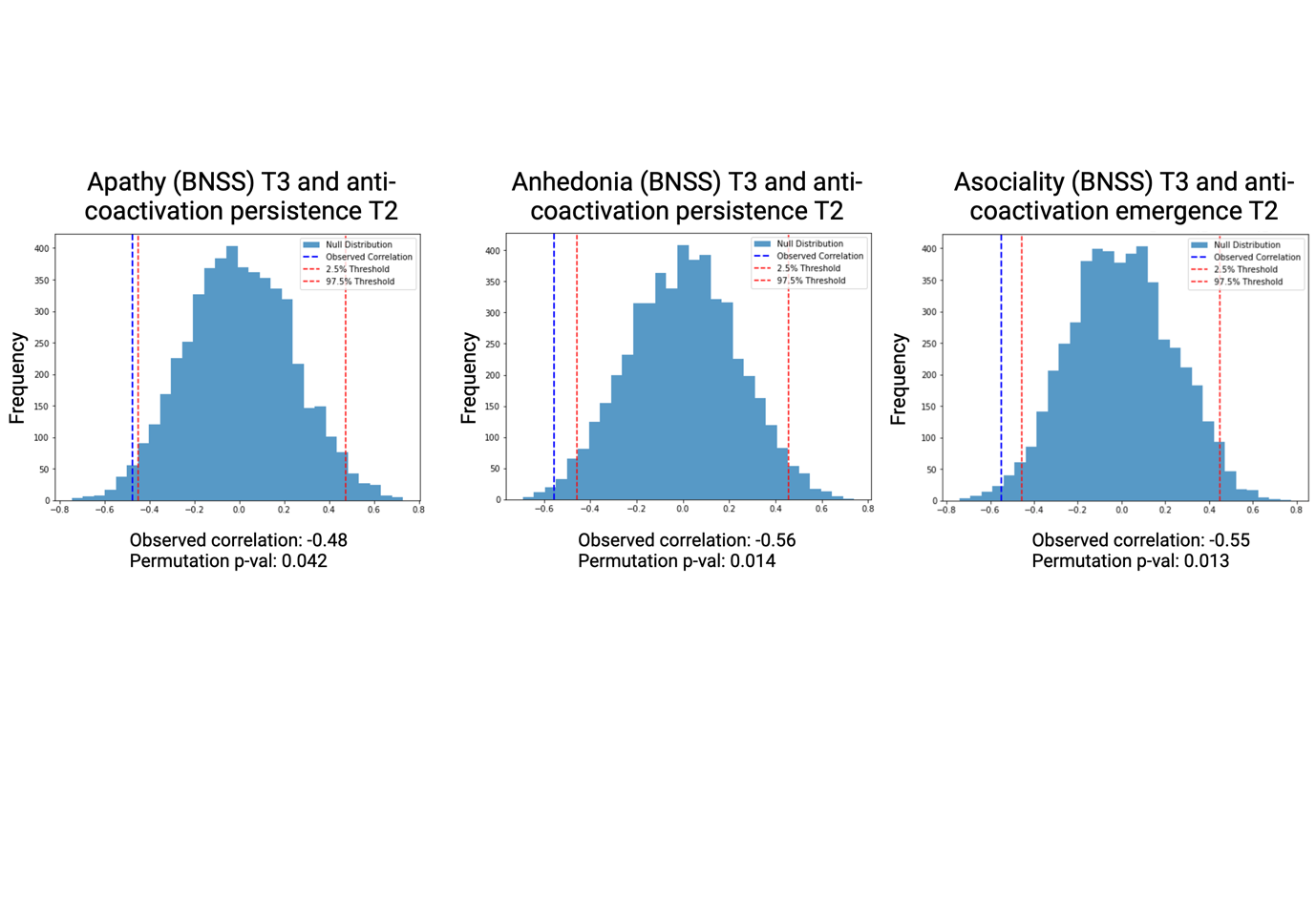

**Figure S11. Main cohort: non-parametric permutation tests between CAP1 metrics at T2 and negative symptoms at T3.**

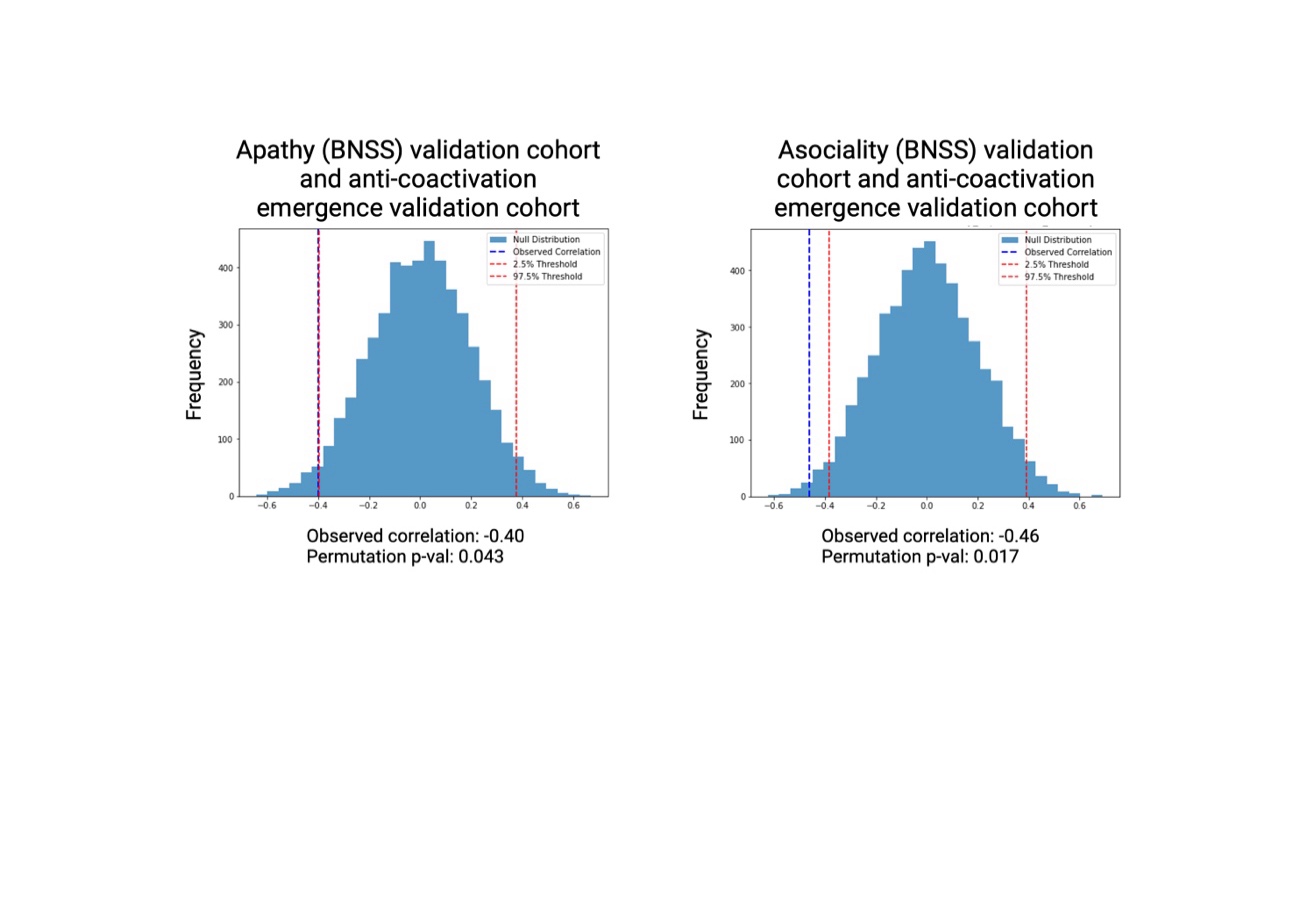

**Figure S12. Validation cohort: non-parametric permutation tests between CAP1 emergence (CAP1 entry frequency) and BNSS apathy and asociality.**
